# COVID-19 reopening strategies at the county level in the face of uncertainty: Multiple Models for Outbreak Decision Support

**DOI:** 10.1101/2020.11.03.20225409

**Authors:** Katriona Shea, Rebecca K. Borchering, William J.M. Probert, Emily Howerton, Tiffany L. Bogich, Shouli Li, Willem G. van Panhuis, Cecile Viboud, Ricardo Aguás, Artur Belov, Sanjana H. Bhargava, Sean Cavany, Joshua C. Chang, Cynthia Chen, Jinghui Chen, Shi Chen, YangQuan Chen, Lauren M. Childs, Carson C. Chow, Isabel Crooker, Sara Y. Del Valle, Guido España, Geoffrey Fairchild, Richard C. Gerkin, Timothy C. Germann, Quanquan Gu, Xiangyang Guan, Lihong Guo, Gregory R. Hart, Thomas J. Hladish, Nathaniel Hupert, Daniel Janies, Cliff C. Kerr, Daniel J. Klein, Eili Klein, Gary Lin, Carrie Manore, Lauren Ancel Meyers, John Mittler, Kunpeng Mu, Rafael C. Núñez, Rachel Oidtman, Remy Pasco, Ana Pastore y Piontti, Rajib Paul, Carl A. B. Pearson, Dianela R. Perdomo, T Alex Perkins, Kelly Pierce, Alexander N. Pillai, Rosalyn Cherie Rael, Katherine Rosenfeld, Chrysm Watson Ross, Julie A. Spencer, Arlin B. Stoltzfus, Kok Ben Toh, Shashaank Vattikuti, Alessandro Vespignani, Lingxiao Wang, Lisa White, Pan Xu, Yupeng Yang, Osman N. Yogurtcu, Weitong Zhang, Yanting Zhao, Difan Zou, Matthew Ferrari, David Pannell, Michael Tildesley, Jack Seifarth, Elyse Johnson, Matthew Biggerstaff, Michael Johansson, Rachel B. Slayton, John Levander, Jeff Stazer, Jessica Salerno, Michael C. Runge

## Abstract

Policymakers make decisions about COVID-19 management in the face of considerable uncertainty. We convened multiple modeling teams to evaluate reopening strategies for a mid-sized county in the United States, in a novel process designed to fully express scientific uncertainty while reducing linguistic uncertainty and cognitive biases. For the scenarios considered, the consensus from 17 distinct models was that a second outbreak will occur within 6 months of reopening, unless schools and non-essential workplaces remain closed. Up to half the population could be infected with full workplace reopening; non-essential business closures reduced median cumulative infections by 82%. Intermediate reopening interventions identified no win-win situations; there was a trade-off between public health outcomes and duration of workplace closures. Aggregate results captured twice the uncertainty of individual models, providing a more complete expression of risk for decision-making purposes.

## Main text

Uncertainty is pervasive during any emerging infectious disease outbreak. There is limited scientific understanding about epidemiological processes, public health and economic goals may be varied, unclear, conflicting, or not stated at all, and the potential effects of possible interventions are uncertain given the novel circumstances. As illustrated by recent outbreaks of Ebola and Zika viruses, and the COVID-19 pandemic, the complexity of an outbreak motivates quantitative modeling, but a profusion of models often produces conflicting forecasts, projections and intervention recommendations (Thomson et al. 2006, Li et al. 2017, Viboud et al. 2018, Carlson et al. 2018, Kobres et al. 2019, Ray et al. submitted). This poses a challenge for decision makers. To support sound, evidence-based decision making, we believe it is critical to develop an efficient framework for collaborative modeling and for synthesizing results and recommendations from ensemble modeling efforts (Tetlock et al. 2014, den Boon et al. 2019).

We previously proposed a method to harness the power of multiple independent research groups and models (Shea et al. 2020) by drawing from tools in decision analysis (Gregory et al. 2012), expert elicitation (Murphy et al. 1998, Burgman 2015, Mukherjee 2015, Tetlock and Gardner 2016), and model aggregation for decision making (Probert et al. 2016, Li et al. 2017). Our approach is designed to reduce unwanted cognitive biases and linguistic uncertainty (e.g., about the interpretation of the problem setting), while characterizing and preserving genuine scientific uncertainty (e.g., about epidemiological processes or parameters, or intervention efficacy, given limited data) that is relevant to policy development and decision making. In this framework, insights can be shared across modeling groups to inform the collective projections, while retaining the perspective of individual groups. The process involves multiple steps (Fig. 1), including two rounds of modeling with an intervening structured discussion to eliminate unwanted biases and uncertainty (including semantic or linguistic uncertainty), increase consistency in modeling of interventions, share critical insights, and generate a comprehensive picture of relevant uncertainty (loop B in Fig. 1). The projections from the second round of modeling are then used to generate aggregate results under different interventions that encapsulate scientific uncertainty about epidemiological processes and management interventions (Li et al. 2019). We stress that this process is designed primarily to inform decision making, rather than to provide quantitative projections of epidemic trajectory (as in ongoing forecasting challenges; Ray et al. submitted), though such results are also obtained. The multi-model, multi-step process is expected to generate better calibrated projections than individual models. That is, the aggregate distributional forecast will be more consistent with future observations than individual forecasts. More importantly, this process is also expected to produce more robust insights about the ranking of intervention options that improve management outcomes. The COVID-19 pandemic offers a unique opportunity to apply this structured framework.

**Fig. 1:**
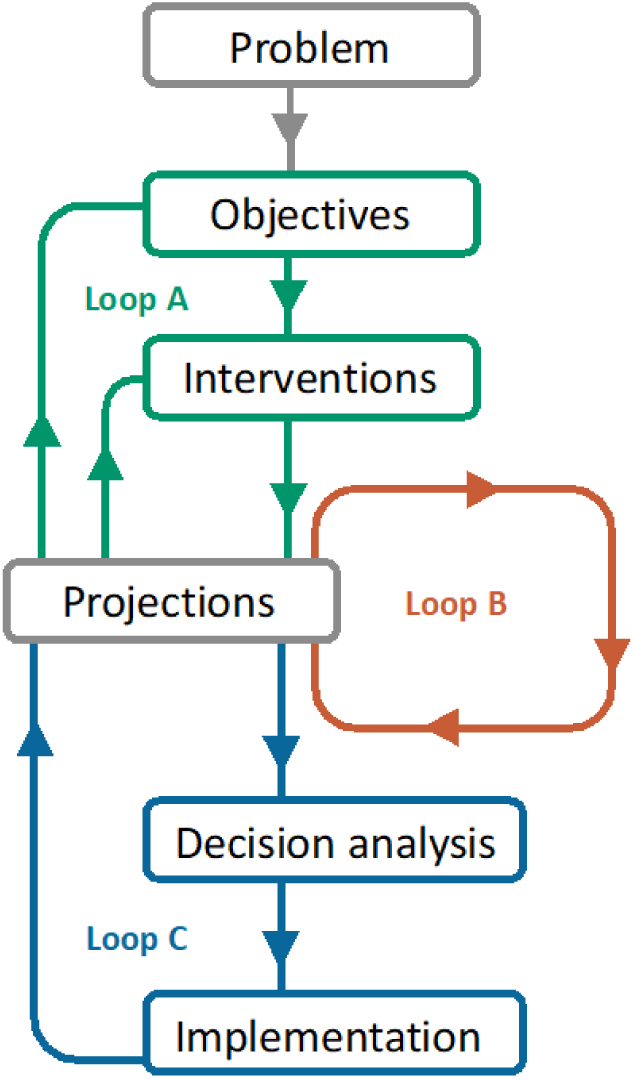
**Multiple Models for Outbreak Decision Support (MMODS) framework,** specifically for the elicitation in this project. The **Problem** is the decision context faced by state and local officials regarding local guidance and regulations concerning the operation of non-essential workplaces, in the face of the COVID-19 pandemic during the period May 15 to November 15, 2020. The 5 **Objectives** addressed were to minimize: (1) cumulative infected individuals, (2) cumulative COVID-related deaths, (3) peak hospitalizations, (4) probability of a new local outbreak (more than 10 new reported cases/day), and (5) total days workplaces closed, all over the period May 15 to November 15. The four **Interventions** focused on strategies for re-opening non-essential workplaces, while assuming all schools remaining closed, between May 15 and November 15, 2020: (1) continue with current non-essential workplace closures, (2) open non-essential workplaces when the number of new daily cases is at 5% of peak, (3) open non-essential workplaces 2 weeks after peak, and (4) immediately relax all current restrictions on non-essential workplaces. Loop B coordinates modeling groups to reduce bias and linguistic uncertainty. First, loop B involves independent (round 1) model **Projections** of all objective-interaction combinations. A structured, facilitated group discussion reduces unwanted uncertainty and also prompts information on additional sources of data used, methods used to incorporate uncertainty, and assumptions made by individual groups, so that the whole collaborative can improve their models. Retention of the remaining model differences allows for a more comprehensive expression of legitimate scientific uncertainty; consensus is not required. Modelling groups then provide updated (round 2) model projections. Loop A provides an opportunity for model groups to interact with decision makers to clarify or update objectives or interventions, i.e., to reduce linguistic uncertainty. **Decision Analysis** is used to aggregate and analyze the model outputs to rank interventions. If decisions are implemented, then there is also an opportunity for modeling teams to learn from **Implementation** data and results (loop C).

Based on this framework, we launched the Multiple Models for Outbreak Decision Support (MMODS) project to guide COVID-19 management in the United States, focusing on a generic mid-sized county of approximately 100,000 people that experienced a small outbreak in late April and early May. Control of COVID-19 in such populations has received relatively little attention but is relevant to decisions faced by state and local officials. We solicited participation from multiple modeling groups via the Models of Infectious Disease Agent Study (MIDAS) network and via existing COVID-19 modeling collaborations with the U.S. Centers for Disease Control and Prevention (CDC) to project outcomes for five objectives related to SARS-CoV-2 incidence, hospital resources, and local workplace and school restrictions over a 6-month period (see Materials and Methods for details). We presented information for a generic, mid-sized county with age structure representative of the U.S. population, that pre-emptively initiated, and adhered to, stringent social distancing guidelines (i.e., full stay-at-home orders with workplace and school closures) until May 15, 2020 (so that the 6-month prediction period ran from May 15-November 15, 2020). We provided the modeling groups with baseline epidemiological and intervention information for the county (see Supplemental Material [SM] File 2 containing provided data); some groups incorporated additional data (see SM Table 1). We considered four possible interventions that mirror the responses of different countries, states, and counties to COVID-19. The four interventions addressed decreasingly stringent non-essential workplace re-openings while assuming schools remained closed, and consisted of (1) closure throughout the 6-month prediction period (“closed” intervention), (2) re-opening when cases decline below 5 percent of the peak daily caseload, (3) re-opening two weeks after peak daily caseload, and (4) immediate re-opening (on May 15, “open” intervention). Each modeling group provided a probability distribution of health and economic outcomes for each intervention, from which aggregate results were generated. After the first round of projections, the aggregate results and anonymized individual results were shared with all groups and a discussion was held to clarify terminology, share insights, and discuss differences. Following the discussion, the groups provided updated projections, from which the final results were generated.

### Aggregate results anticipate outbreaks for any level of reopening

Sixteen modeling groups participated in this study, contributing 17 distinct models with a variety of structures and assumptions (see Materials and Methods, and SM Table 1) and two rounds of projections. Results are presented using projections from the second round only. The aggregate projections showed a consistent ranking of intervention performance (SM Figs. 1 and 8 and SM Video 1) across the four public health outcomes (cumulative infections, cumulative deaths, peak hospitalization, and probability of an outbreak), and a strong trade-off between public health and economic outcomes (number of days with non-essential workplaces closed over the 6-month period, Fig. 2). For all of the public health-related outcomes, the best intervention was to keep non-essential workplaces closed for the duration of the period investigated. Reopening at 5% of the peak and reopening 2 weeks after the peak were ranked second and third, respectively. Opening fully and immediately led to the greatest public health burden. Keeping restrictive measures in place for 6 months reduced median cumulative infections by 82%, from a median of 48,100 (48.1% of the county population) in the open intervention to 8,527 in the closed intervention; the 5-percent and 2-week interventions reduced the cumulative infection by 66% and 46%, respectively, relative to the open intervention. The reduction in cumulative deaths followed a similar pattern (Fig. 2). Peak hospitalizations ranked the interventions in the same order as cumulative cases and deaths, but the largest decrease was achieved when going from the open intervention to the 2-week intervention.

**Fig. 2:**
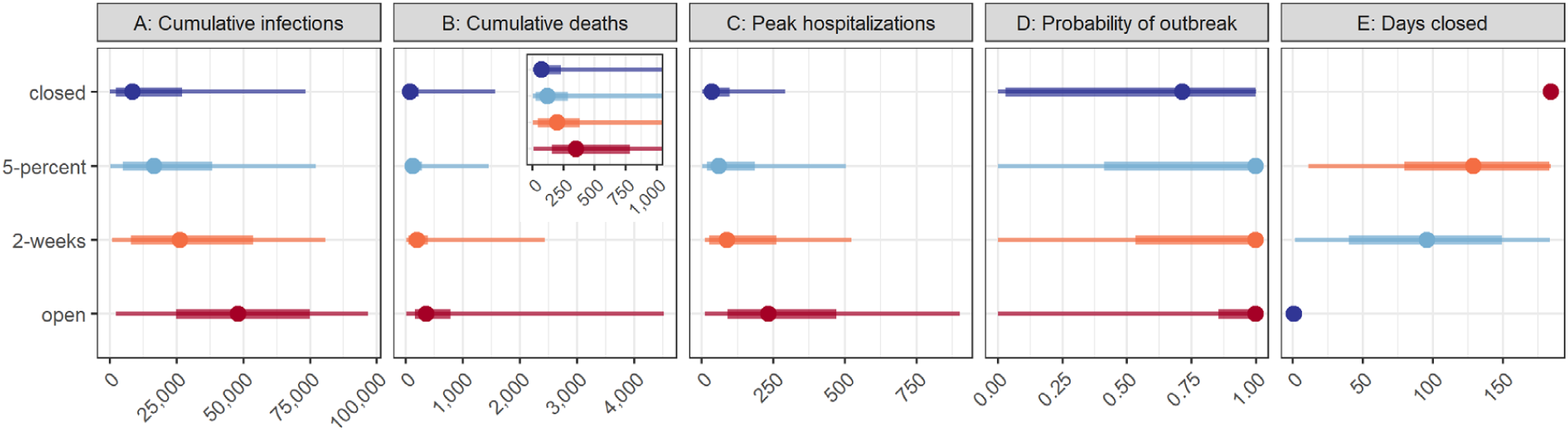
Aggregate distribution for target objective and intervention scenario pairs of the 17 models. Median, 50% prediction interval (PI), and 90% PI are indicated as points, thick lines, and thin lines respectively. The aggregate distribution was calculated as the weighted average of the individual cumulative distribution functions. Colors denote ranking of each intervention for a single objective, where dark blue signifies the lowest value (best performance) and dark red signifies the highest value (worst performance). The five panels show the results for: A) cumulative infections (rather than reported cases) between May 15 and November 15; B) cumulative deaths due to COVID-19 over the same period, with an inset displaying the results for a smaller range of values, beginning with zero and containing the 50% prediction intervals; C) the peak number of hospitalizations over the same period; D) the probability of an outbreak of greater than 10 new cases per day after May 15; and E) the number of days that non-essential workplaces are closed between May 15 and November 15. The interventions include: “closed”, workplace closure throughout the 6-month period; “5-percent”, non-essential workplace re-opening when cases decline below 5% of the peak caseload; “2-weeks”, non-essential workplace re-opening two weeks after the peak; and “open”, immediate re-opening of all workplaces. The setting is a generic US county of 100,000 people that has experienced 180 reported cases and 6 deaths as of May 15, 2020; all schools are assumed to be closed throughout the period.

We asked the modeling groups to estimate the probability of an outbreak after May 15 (defined as a 7-day moving average of reported cases greater than 10 per day). Even under the fully closed intervention, the probability of an outbreak was high (aggregate median, 71%). The median probability of an outbreak increased to 100% for all other interventions. Even relatively stringent re-opening guidelines were insufficient to guarantee success; complete cessation of community spread of the disease was unlikely even with long-term non-essential workplace closure (although widespread transmission has been pre-empted in some settings, such as New Zealand and Taiwan, with high compliance to a package of social distancing measures and travel restrictions). Either additional stay-at-home orders would be required, or other non-pharmaceutical interventions (e.g., testing, contact tracing and isolation, or wearing masks) or pharmaceutical interventions (e.g., vaccination) would be needed to stop transmission while allowing workplace re-opening.

Our results allowed us to examine trade-offs between economic and public health outcomes; how much economic activity might the decision maker be willing to forgo to gain public health improvements of a given magnitude? The number of days of non-essential workplace closure is a coarse and incomplete measure of short-term economic impact, but it highlights an important trade-off. The ranking of interventions in relation to projected days closed was reversed in comparison to the ranking for the public health outcomes (Fig. 2): under the closed intervention, non-essential workplaces were closed for 184 days (May 15 to November 15); under the 5-percent and 2-week interventions, non-essential workplaces were closed for a median of 129 and 96 days (a reduction of 29% and 48%), respectively. We had hypothesized that the 5-percent intervention might be an attractive alternative relative to remaining closed, permitting a reduction in days closed with little or no difference in public health outcomes. However, cumulative infections, deaths, and peak hospitalizations were further reduced under the closed intervention relative to the 5-percent intervention by 48% (8,527 vs. 16,510), 39% (73 vs. 119), and 40% (36 vs. 60), respectively (Fig. 2). This starkly illustrates the tensions between economic and public health goals seen worldwide and suggests that strategies that only consider the timing of re-opening, or focus on a single type of intervention, may not be nuanced enough to manage these trade-offs.

### Benefits of the structured MMODS process

The MMODS process outlined here is focused on decision outcomes and, unless a rapid decision is required (necessitating the interim use of round 1 results), only round 2 results are considered. However, as this is a new approach to ensemble decision making for outbreak control (Shea et al. 2020), results about the changes between the first and second rounds are pertinent. The group discussion after the first round identified numerous sources of linguistic uncertainty arising from different interpretations of the objectives and the nature of interventions (for example, the definition of ‘death’; see SM on Resolution of Linguistic Uncertainty for details). Clear guidelines developed during and after the group meeting removed this uncertainty from round 2 projections, improving the comparability of intervention rankings across models. The discussion also motivated the modification of one reopening intervention (Fig.1 loop A; see Materials and Methods). The discussion highlighted additional sources of available data, shared critical insights with all groups, and encouraged a broader expression of scientific uncertainty (including a discussion of different methods used for incorporating such uncertainty), all while maintaining anonymity of results to avoid pressure to conform where true scientific disagreement persisted. As linguistic uncertainty was decreased at the same time as modeling groups were encouraged to more fully express their remaining uncertainty, there is no expectation of directionality in the relative magnitude of uncertainty expressed by models in the two rounds of projections. However, projections of days closed in the two extreme (open and closed) interventions were highly variable in round 1, but are entirely consistent in round 2 (compare Fig. 3 for open and closed interventions, top and bottom panels in column 5, with SM Fig. 9), demonstrating the complete removal of linguistic uncertainty that would have confounded results.

**Fig. 3:**
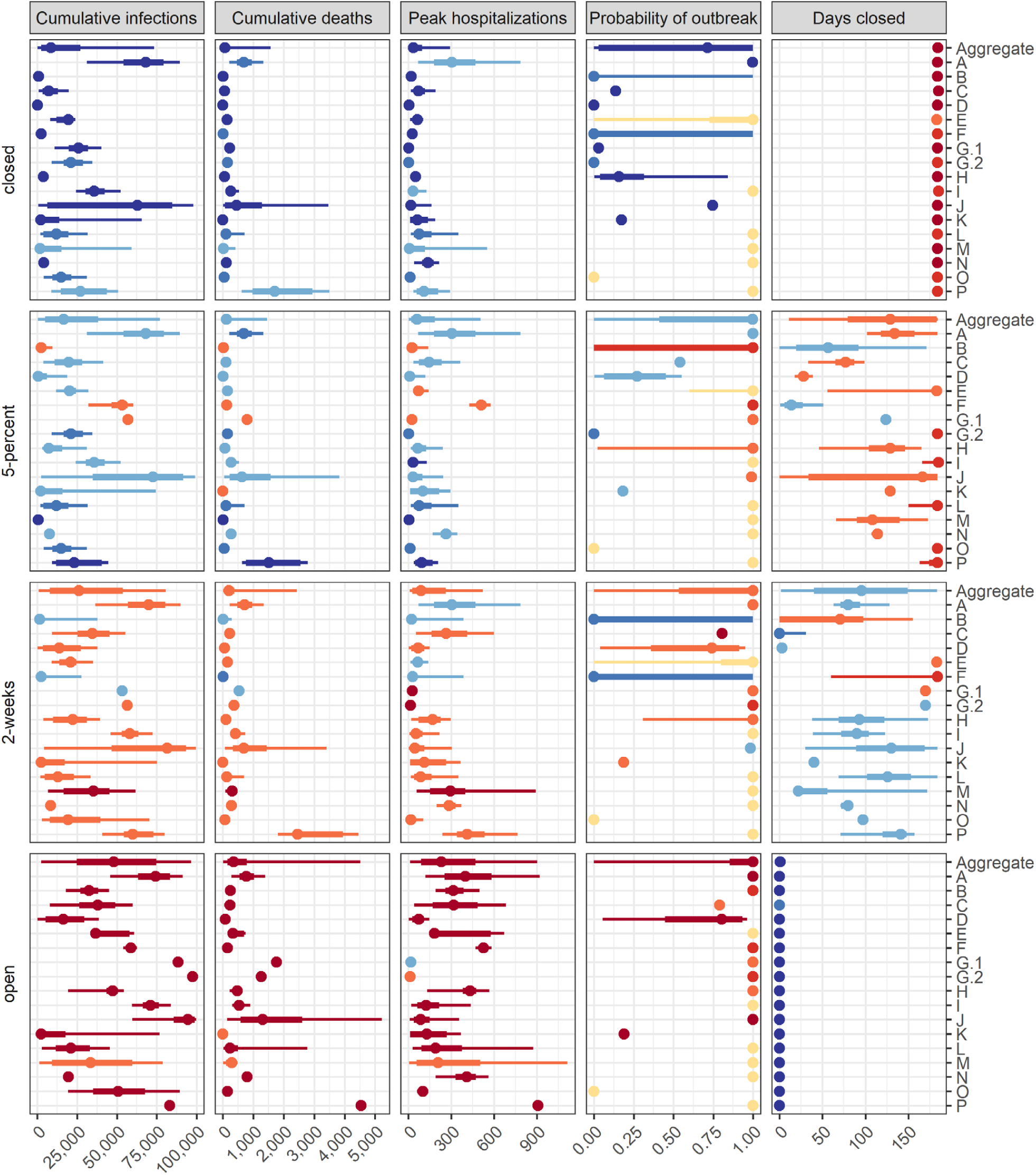
Individual model results for each objective and intervention scenario pair. Median, 50% prediction interval (PI), and 90% PI are indicated as points, thick lines, and thin lines respectively. Colors denote ranking of each intervention by model for a single objective, where dark blue signifies the lowest value (best performance) and dark red signifies the highest value (worst performance). Ties in ranks are colored as intermediate values. Ties between ranks 1 and 2 and ranks 3 and 4 are shown as an intermediate blue and red, respectively; yellow indicates a tie in ranks across all interventions. Each group was assigned a random, unique identification letter that is specified on the vertical axis.

### Individual models are consistent in ranking of interventions, but projections are variable in magnitude and uncertainty

The rankings of interventions from the individual models are generally consistent with each other and with the aggregate results (Fig. 3 and SM Video 1); however, there are some interesting differences. Three models ranked the closed and 5-percent interventions as identical for several metrics; 6 groups reported that for at least some simulations the 5-percent reopening criterion was never met in the 6-month period. Two models ranked the 5-percent intervention as better than the closed intervention based on the medians of health outcomes; both models had wide priors on parameters governing compliance with interventions. Three of the 17 models ranked the 5-percent intervention worse than the 2-week intervention for public health measures (Fig. 4A i-iii), a result that was driven by different timing in the triggering of re-opening (Fig. 4A iv). Another notable result is that the ranking of the 2-week intervention for the peak hospitalization metric spanned the gamut from worst rank (in submission M) to first-tied rank (in submission A) (Fig. 3). Otherwise, rankings were remarkably consistent overall.

**Fig. 4:**
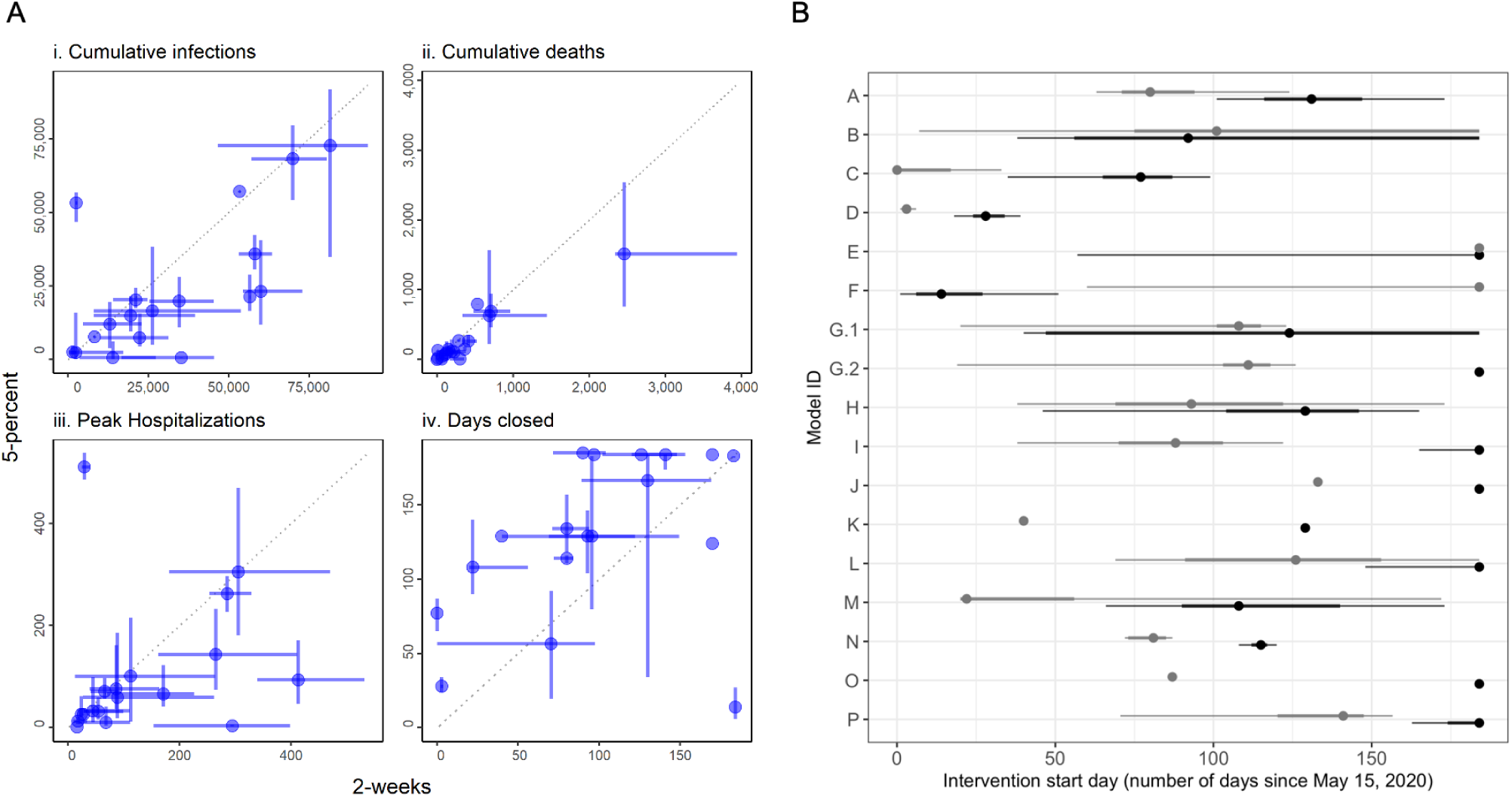
Comparison between the 2-week and 5-percent interventions. **A)** Medians (points) and 50% PIs (lines) displayed pairwise by intervention and for the following objectives: i) cumulative infections, ii) cumulative deaths, iii) peak hospitalizations, and iv) days closed for each model. **B)** Comparison of intervention start dates for 2-week (grey) vs. 5-percent (black) interventions for each model, where the start date is computed as the number of days from May 15 until the intervention is enacted. Intervention start times of 184 days indicate that the intervention was never triggered in that model. All plots display median (points) and 5 ^th^ to 95 ^th^ quantiles (lines) for each intervention. The 2-week intervention trigger to open is the first day for which the 7-day trailing moving average of the number of new daily reported cases has been lower than the maximum for at least 14 days, and has shown a day-to-day decline in smoothed case data for at least 10 of the last 14 days (or, there have been 7 days without any new cases). The 5-percent intervention trigger to open is the first day for which the 7-day trailing moving average of the number of new daily reported cases drops below 5% of the maximum.

Importantly, the individual models display considerable variation in terms of the magnitude and uncertainty of projections (Fig. 3). Reliance on a single model, rather than an ensemble, is inherently less reliable for providing insights on the magnitude of differences between interventions, even if the ranking of interventions is relatively robust (SM Fig. 1). In fact, there are numerous examples where even the 90% prediction intervals (PIs) do not overlap for different models in the same intervention scenario (for example, see cumulative infections for the open intervention, where all models rank the intervention as worst, but the range of the population infected nearly covers 0 to 100%). The differences in model structure, parameterization, and assumptions that the groups were asked to provide did not explain differences in the results (see Materials and Methods and SM Tables 1, 2, Figs. 15, 16); such an evaluation, facilitated by a thorough model description checklist, would be impossible with individual experts. This pattern of consistent rankings across models despite a wide variation in projected outcomes was also seen for Ebola (Li et al. 2017). Estimating the relative difference in an outcome across different courses of action (a decision-making task) is almost always more straightforward than estimating an absolute value (a prediction task), as the former simply involves a rearrangement of ordinal rankings, while the latter has to account for many more factors.

### Aggregate results provide an integrated expression of uncertainty

It is challenging for any individual model alone to fully account for uncertainty. The aggregate results provided a more comprehensive measure of uncertainty by integrating over the individual team assumptions about disease dynamics, population behavior, public health surveillance, and the effectiveness of interventions. Deploying multiple models in parallel also speeds up the process of exploring relevant uncertainty. Individual models tended to capture less than 50% of the uncertainty of the aggregate (as measured by the relative interquartile range, IQR, Fig. 5). As a result, individual models were generally more confident than the ensemble, echoing findings from studies of expert judgment that individual experts tend to be over-confident (Teigen and Jørgensen 2005). The importance of a well-calibrated expression of uncertainty in projections can be seen in the context of hospital planning, and the risk tolerance a hospital administrator needs to contemplate.

**Fig. 5:**
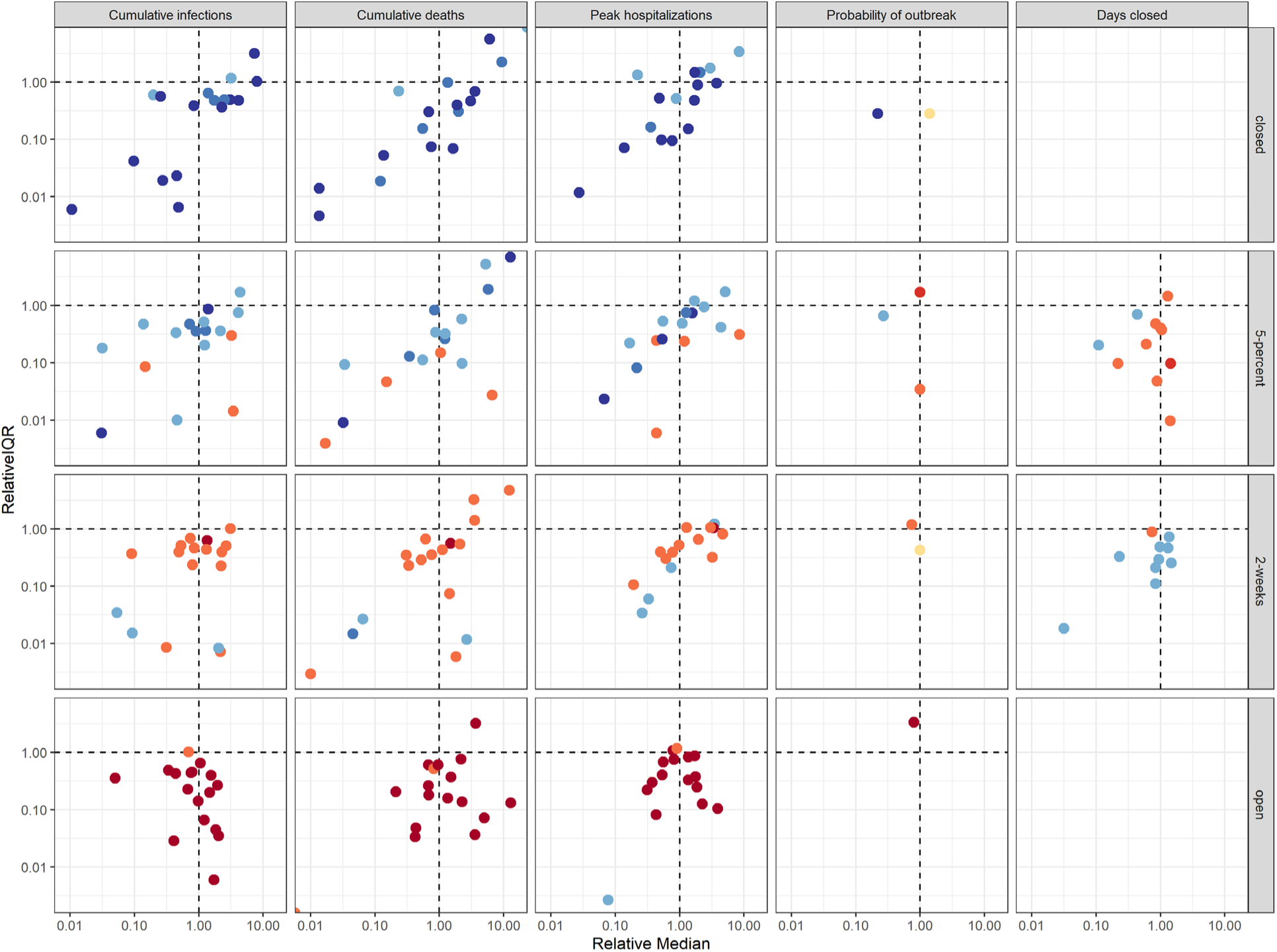
Comparison of individual model results to aggregate results. The y-axis shows the relative interquartile range (IQR)—the ratio of an individual model’s IQR to the aggregate IQR. The x-axis shows the ratio of an individual model’s median to the aggregate median. Both axes are presented on a log scale. Colors denote ranking of each intervention by models, where dark blue signifies the lowest value (best performance) and dark red signifies the highest value (worst performance). Ties between ranks 1 and 2 and ranks 3 and 4 are shown as an intermediate blue and red, respectively; yellow indicates a tie in ranks across all interventions.

For example, if the county has 200 hospital beds, while median peak hospitalization is comparable for the remain-closed and 2-week interventions, the 2-week intervention was three times as likely to exceed capacity as the remain-closed intervention (34% vs. 11% chance of exceedance based on the aggregate results, Fig. 2). These risk estimates allow the administrator to gauge how much to prepare for exceedance. If a local official or hospital administrator, however, had to rely on only a single model to estimate the exceedance risk, they may mis-estimate the risk and possibly over- or under-prepare.

### Model projections are comparable with real-world data

We identified 66 mid-sized (90,000 to 110,000 people) US counties that approximated the profile of the setting presented to the modeling groups and that have implemented and followed a closed intervention (e.g., a stay-at-home order) through October 15, 2020 (Dong et al. 2020) (see SM section: Comparison of county data with aggregate model results and MMODS code repository). The distribution of reported deaths due to COVID-19 in the closed counties (median 31; 50% IQR, 16 to 56) was comparable to the aggregate projection of total deaths (both reported and undetected, median 73; 50% IQR, 12 to 228). The confidence intervals for projected deaths were wider than the observed distributions, which is expected as the observations represent a subset of the possible paths that the outbreak might have taken (see SM Figs. 1, 3, 13 and text in SM section: Comparison of county data with aggregate model results). This analysis will be updated and posted to the SM after November 15, 2020 (once the prediction period is complete). If the MMODS decision process were used to guide decision making (Fig. 1, loop C), there is the opportunity to weight models in the ensemble as we learn about their performance, or for modeling teams to learn from data and improve their models in an adaptive management framework (Shea et al. 2014, Probert et al. 2018, Shea et al. 2020a, b).

## Discussion

The abundance of uncertainty that accompanies pathogen emergence presents a uniquely difficult challenge for public health decision making. The multi-stage, multi-model process revealed some important epidemiological and policy insights about county re-openings. The aggregate results, and most individual models, ranked the interventions consistently for any given objective. While more stringent reopening rules generally performed better, public health strategies designed only around one-time re-opening guidelines were inadequate to control the COVID-19 epidemic at the county level, as reflected in the resurgence of COVID-19 over the summer of 2020 in the United States. Our results reinforce the importance of coupling these strategies with other pharmaceutical, non-pharmaceutical, and behavioral interventions (e.g. vaccine deployment, mask usage, and social distancing).

The descriptions of the objectives and interventions for this elicitation were motivated by public discussions and guidance issued by federal and state governments in April 2020. However, concepts presented in colloquial language can be difficult to precisely define mathematically. All groups found that the initial wording in the guidance provided was difficult to interpret and model, suggesting it could invite considerable discretion in implementation. For example, the wording on reopening ‘2 weeks after peak’ engendered considerable confusion in the first round of modeling. How is a peak defined? Is it in reported deaths or cases? Is it measured on a daily or a moving-average basis? Likewise, how should a model determine whether 2 weeks have passed since the peak? A continuous monotonic decline was never seen; should a moving average be used? And, if so, for 7 or 14 days? We provided a clear definition of such terms for the second round of projections (see Materials and Methods for definitions, and SM on Resolution of Linguistic Uncertainty for discussion). Even so, there was still considerable variation across modeling groups in how these openings were triggered, in part because the triggering events were sensitive to how daily variation in the projections was handled (Fig. 4B). More precisely worded guidelines, and crucially, clear lines of communication and open collaboration between decision makers and modelers (Fig. 1, loop A), could reduce confusion, and would permit consistent evaluation and application of management interventions. Our process illuminated this issue and created a well-defined and timely opportunity to resolve it, ensuring more comparable results on which to base decisions.

Balancing public health outcomes and economic considerations is an important aspect of pandemic decision making, but needs a more nuanced treatment, particularly on the economic side. If we compare each of our four interventions to a hypothetical “no disease” scenario, we can identify four broad groups of costs from multiple perspectives: (a) financial costs caused by the disease itself (e.g., reduced economic output due to absence from work due either to illness or voluntary isolation, direct and indirect costs of medical treatment, or costs associated with funerals); (b) non-financial costs caused by the disease itself (e.g., mortality, morbidity, long-term health impacts); (c) financial costs caused by the strategic response to the disease (e.g., reduced economic output due to a lockdown, costs of monitoring and enforcing a lockdown or of quarantining incoming travelers) or by individual responses that go beyond local policy; and (d) non-financial costs caused by strategic or personal responses (e.g., mental health challenges due to isolation, forgoing preventive medical care such as routine childhood vaccinations, relationship stressors, reduced access to opportunities for recreation). There may also be economic benefits associated with the mitigation activities. For example, some firms have found that having staff work from home can increase efficiency and reduce operational costs (OECD 2020), which could have ongoing benefits. In principle, the optimal strategy would be that which minimizes the sum of all these costs, less any benefits. Our analysis does not quantify all of these costs and benefits but does provide evidence that could be used as key inputs to a comprehensive economic analysis. Feedback to decision makers from this process may lead to refined or multi-criteria objectives (via loop A in Fig. 1).

Running the models twice, with an intervening discussion, is essential. In our elicitation, we removed different interpretations of terminology that would have confounded the ensemble results. For example, the trade-off between days closed and public health outcomes would have been obscured by linguistic uncertainty surrounding “closure” (see SM Fig. 9). The importance of consultation and open collaboration between decision makers and modelers (Fig. 1, loop A) to clarify objectives and interventions is also strongly supported. Initially it might seem that adding a second step would delay decision-making; this is a valid concern in time-sensitive circumstances. In an emergency, results from round 1 could be used to inform an interim decision. However, in practice, the two-round process generally can be more efficient. Clarification often happens on an *ad hoc* basis anyway as, despite best efforts, it is fundamentally difficult to anticipate everything that groups might interpret differently *a priori* (though it will be possible to learn and streamline across exercises), and uncertainties often arise as models are developed and implemented. Our process is deliberate in explicitly planning for and appropriately managing this process, so that all groups are equally informed and use the same interpretations. Formally building the discussion phase into the modeling and decision-making process manages decision-maker expectations and saves time in the long run, by avoiding multiple, unplanned reassessments. This would be particularly valuable in situations where the same models are used to make repeated decisions. Modeling teams also commented that they found the well-defined structure in Fig. 1 to be valuable. We further note that resolution of unwanted linguistic uncertainty via facilitated group discussions would likely also improve forecasts outside the decision-making setting.

Multiple model approaches such as MMODS come with challenges, not least of which is coordinating multiple, disparate groups’ efforts. Participating modeling groups refocused their efforts on COVID-19 during the past year, and contributed considerable, mostly unfunded, time and effort (see SM Table 1) to participate in this project on a voluntary basis. Not all models were initially structured to address the questions or interventions we considered, and the methods for handling uncertainty were novel for some of the groups. Additionally, we found some trade-offs were necessary. A number of the models (e.g., those with substantial simulation time, or requiring extensive changes to implement new interventions) could not assess the impact of many distinct interventions, so we limited our study to four workplace-related interventions to encourage participation. However, this need not constrain the number of interventions for critical decisions and situations; it is possible to augment computational resources, or different subsets of the contributing models could assess different subsets of interventions. Most of these challenges could be addressed via planning and preparedness efforts to build a community of epidemiological and decision-making experts in anticipation of need (Drake 2020, National Academies of Sciences, Engineering, and Medicine 2020, Rivers et al. 2020). The success of future collaborative efforts will depend on the sustained availability of financial and logistical support, for both the coordination of collaborations, and for the individual modeling groups. Such coordinated efforts could then provide rapid, informed support to decision makers at critical junctures, to enable effective decision making in the face of uncertainty, as well as the opportunity for rapid, focused learning through monitoring and model-weight updating (Shea et al. 2014).

Our project is the first open study to use multiple models, instead of multiple individual experts, in a structured expert elicitation process focused on real-time disease mitigation decisions. As with well-designed expert elicitations, using multiple models produces a more complete description of uncertainty and provides more robust projections to decision makers. While we have drawn from the expert elicitation literature (Runge et al. 2011, Burgman 2015) to design this process, more work is needed to optimize the approach (Shea et al. 2020a). For example, a key question that has not yet been addressed in model elicitations is whether an open call for participation or a curated set of established models produces better results. Both approaches (open participation and careful selection) are used with expert panels, but often rely on different analytical methods. Second, how many models are needed to produce stable and robust results? In individual expert judgment approaches, between 5 and 20 experts are recommended (Rowe and Wright 2001); however, it is not clear if the same guidance applies for models.

We have outlined these methods, and their benefits and challenges, for reopening decisions in mid-sized US counties facing the COVID-19 pandemic. However, this approach can be applied to a wide range of other settings or to other critical decisions, such as: when to reimpose or relax interventions in a sequence; context-dependent state- and country-level interventions (including in low- and middle-income countries where constraints and available resources may differ markedly on a case-by-case basis, e.g., Walker et al. 2020); where best to trial vaccines and drugs; how to prioritize testing; and how to optimize the roll-out of other medical interventions. In all cases, the MMODS approach demonstrated here could reduce unnecessary uncertainty in terminology and interpretation, better characterize the remaining scientific uncertainty, reduce bias and minimize the temptation to rapidly reach premature consensus. This approach can also be used for critical management decisions for endemic diseases, and for elimination and eradication planning, as well as in any non-epidemiological setting where models are used to inform decision making (e.g., Milner-Gulland and Shea 2017). We also stress that our approach encourages an integration of science and policy-making – efforts that are often separated to the detriment of public health outcomes when semantic uncertainties cannot be clarified and may thus interfere with success. Modelers intend their forecasts to ‘inform management decisions,’ yet the common separation of model outputs from the decision context increases the chance of misunderstandings and errors. Continued efforts to foster collaboration and streamline communication between modelers and decision makers, as well as to shift the focus from solely providing projections to evaluating proposed interventions, are essential steps towards effectively leveraging modeling efforts to inform decisions. Such advances will be essential to support real-time decision making on many critical problems.

## Materials and Methods

We solicited participation from multiple modeling groups via the Models of Infectious Disease Agent Study (MIDAS) network and via working groups involving modelers and the U.S. Centers for Disease Control and Prevention (CDC) facilitated by the MIDAS Coordination Center at the end of May 2020. Information about the collaboration opportunity was communicated via conference calls and listservs. This activity was reviewed by CDC and was conducted consistent with applicable federal law and CDC policy (45 C.F.R. part 46, 21 C.F.R. part 56; 42 U.S.C. §241(d); 5 U.S.C. §552a; 44 U.S.C. §3501 et seq). Full information for the elicitation, including the setting, epidemiological data, and intervention descriptors was posted at a dedicated website at https://midasnetwork.us/mmods/ (including daily reported cases, deaths, mobility, and testing data, State of Emergency and stay-at-home orders, and age structure). The initial conditions for the forecasts included cumulative cases and deaths within the county on a daily basis from January 22 to May 15, 2020. As of May 15, the county had recorded 180 confirmed cases and 6 deaths due to COVID-19. Groups were also permitted to incorporate additional data sets, as they saw fit (e.g., national data on hospital, Intensive Care Unit and ventilator availability, household size data, and work, school, community, and home mixing data). We asked the modeling groups to assume that travel restrictions remained in place throughout (so that there was no international importation of cases and domestic importations were limited), and that there was no local contact tracing or isolation of infected individuals. We did not specify guidelines regarding mask use but specified that schools would remain closed through November 15 (just prior to the start of peak flu season). First round results were due on June 15, 2020 and the group discussion of preliminary results took place on June 24, 2020. Second round model results were due 12 July, 2020 and preliminary analyses of second round results were reported to the modeling groups and others on July 17, 2020.

The five objective metrics were: (1) cumulative number of infected individuals (May 15 to November 15); (2) cumulative number of COVID-related deaths over the same period; (3) peak hospitalizations during the period May 15 to November 15; (4) probability of a new local outbreak (more than 10 new reported cases per day); and (5) total number of days workplaces closed. The four interventions focused on strategies for re-opening non-essential workplaces, while assuming all involved schools remaining closed: (1) continue with current non-essential workplace closures at least through November 15 (“closed intervention”), (2) open non-essential workplaces when the number of new daily reported cases is at 5% of peak (“5-percent intervention”), (3) open non-essential workplaces two weeks after peak (“2-week intervention”), and (4) immediately relax all current restrictions on non-essential workplaces on May 15 (“open intervention”).

All objective-intervention combinations were assessed for feasibility in an in-house model prior to developing the elicitation. However, intervention (2), which was set at 1% in round 1, was identified as too restrictive (i.e., the condition was never met) by several models during the discussion, and therefore changed accordingly (i.e., modeling groups provided feedback on interventions as anticipated in Fig. 1 loop A). In a single-round elicitation such a situation would have effectively reduced the number of interventions examined overall, as the 1% trigger was essentially congruent with the fully-closed intervention.

For the 5-percent and 2-week interventions (interventions 2 and 3), following the resolution of linguistic uncertainty in the discussion (see SM: Resolution of linguistic uncertainty), we asked all groups to use the same metric and method for calculating the peak, acknowledging that this is only one of several metrics and methods that could be used to determine the peak. We chose a definition that could be implemented by a decision maker (as opposed to an omniscient approach). For both the 2-week and 5-percent interventions, all teams used the 7-day trailing moving average of the number of new daily reported cases (as opposed to all infections, which may or may not result in reported cases); the moving average smooths out noise due to reporting and low population size. Peak is then defined as the maximum 7-day moving average of daily reported cases. The trigger to open for the 2-week intervention is the first day for which the 7-day trailing moving average has been lower than the maximum for at least 14 days, and has shown a day-to-day decline in smoothed case data for at least 10 of the last 14 days (or, there have been 7 days without any new cases). The trigger to open for the 5-percent intervention is the first day for which the 7-day trailing moving average of the number of new daily reported cases drops below 5% of the peak after May 15 ^th^. Note that the peak that triggers a 2-week intervention may not be the same peak that triggers a 5-percent intervention (e.g., if there is a second peak that is larger than the first one).

Each group completed an extensive checklist (see SM File 3 and SM Table 1) for each round, to document a wide range of information on model structure and parameterization, the efficacy of interventions, additional setting information, assumptions and the associated uncertainty, as well as other sources of stochasticity (see SM Tables 1, 2 and SM Figs 15 to 17). Of the 17 models contributed by 16 scientific research groups, 10 were compartmental, 5 individual-based, 3 spatially explicit, 1 neural network, and 1 fractional order model. Eleven models included age-structure explicitly for some model components. Models handled uncertainty using different methods for different components (e.g., expert judgment, likelihood-based, or simulation methods). As part of the submission checklist, model groups were asked to provide an estimate of the number of Full Time Equivalent (FTE) hours allocated to their modelling effort, so we could assess the resources required to undertake such a multi-model effort. Modeling groups allocated an estimated 64 median FTE hours (Q1: 40 FTE hours, Q3: 100 FTE hours, Max: 1000 FTE hours). No groups dropped out between rounds. One group did not submit a model for round 1, but participated in the discussion and submitted to round 2.

We requested 100 quantiles for each model-objective-intervention combination such that tail probabilities for the 2nd and 98th quantiles were relatively stable (i.e., we requested the probability distribution for each outcome for each intervention, via the cumulative distribution function (CDF) in 100 quantiles). Requesting quantiles (rather than, for example, epidemiological curves) enables all types of different models to participate and allows a better expression of uncertainty for decision making. Collecting 100 quantiles allows the tail probabilities to be estimated. We deliberately did not request information on the correlation structure between interventions within a model, as not all models were equipped to provide results for the same initial conditions or seed values.

Submissions were received by the MIDAS Coordination Center through the MMODS website, verified for format compliance, transformed into a consistent format for analysis, and deposited in an internal project GitHub repository.

Aggregate results were produced by taking a weighted average of the individual cumulative distribution functions (SM Fig. 7); this provides critical information about the mean as well as higher order moments. Each group received equal weight in the aggregate results. For research groups submitting more than one model, the group weight was divided equally among their models (one group submitted two models). Based on the detailed checklist information, we can explicitly document the differences between models and the CDFs reflect the full degree of uncertainty considered. This model-based approach presents two advantages over human experts, who generally provide 3 to 5 quantiles at most, and generally do not document explicit differences in their thought processes that might generate different rankings (Burgman 2015). While it is impossible to do a full analysis of every difference between all the models, we explored multiple potential correlates of model result rankings and magnitudes. Ancillary information was examined to assess whether model assumptions (e.g., model structure, assumptions about importations, etc.) predicted ranks or magnitudes of projections, or various other aspects of epidemic dynamics (e.g., projected number of people who are susceptible on November 15): nothing obvious emerged (see SM: Checklist data and SM Table 1). There also was no obvious uncertainty that would reverse the choice of optimal intervention (e.g., a factor whose inclusion or exclusion leads to different rankings); had such a factor arisen, this would be a top priority for research to improve decision-making outcomes. We also document where differences in magnitude between two strategies are not large; this is only the case for individual models, but not in the aggregate. In such cases, the decision maker may have flexibility of choice, and may choose to weigh other considerations (such as costs) that have not been explicitly included in the models. Magnitude was not our primary interest, but is important in determining whether the overall benefits of an intervention are sufficient to outweigh the overall costs.

## Data Availability

This is a modeling paper; a Github repository for the code and estimates will be provided.

## Acknowledgements

The MMODS team acknowledges Bryan Grenfell and Ottar Bjørnstad. Individual modeling team acknowledgements are listed in the SM.

## USGS Disclaimer

Any use of trade, firm, or product names is for descriptive purposes only and does not imply endorsement by the U.S. Government.

## CDC Disclaimer

The findings and conclusions of this report are those of the author(s) and do not necessarily represent the official position of the Centers for Disease Control and Prevention.

## NIST Disclaimer

These opinions, recommendations, findings, and conclusions do not necessarily reflect the views or policies of the National Institute of Standards and Technology or the United States Government.

## Supplemental Material (SM)

**Supplemental Figures, Tables, Model Descriptions, and other materials**

### Supplementary Material TOC

- Setting data and original elicitation
- Submission checklist
- Anonymized results from round 2 (SM Figs 1 - 8)
- Video showing ranking of round 2 results across models (SM Video 1)
- Resolution of linguistic uncertainty in structured discussion between rounds 1 and 2 (SM Figs 9 - 12)
- Comparison with U.S. county data (SM Figs 13 - 14)
- Checklist data, including contributed model descriptions and funding acknowledgments (SM Tables 1-2, SM Figs 15 - 17)
- MMODS code

### Supplementary Material: Anonymized results from round 2 (Figs. S1 to S8 and Video 1)

**SM Fig 1:**
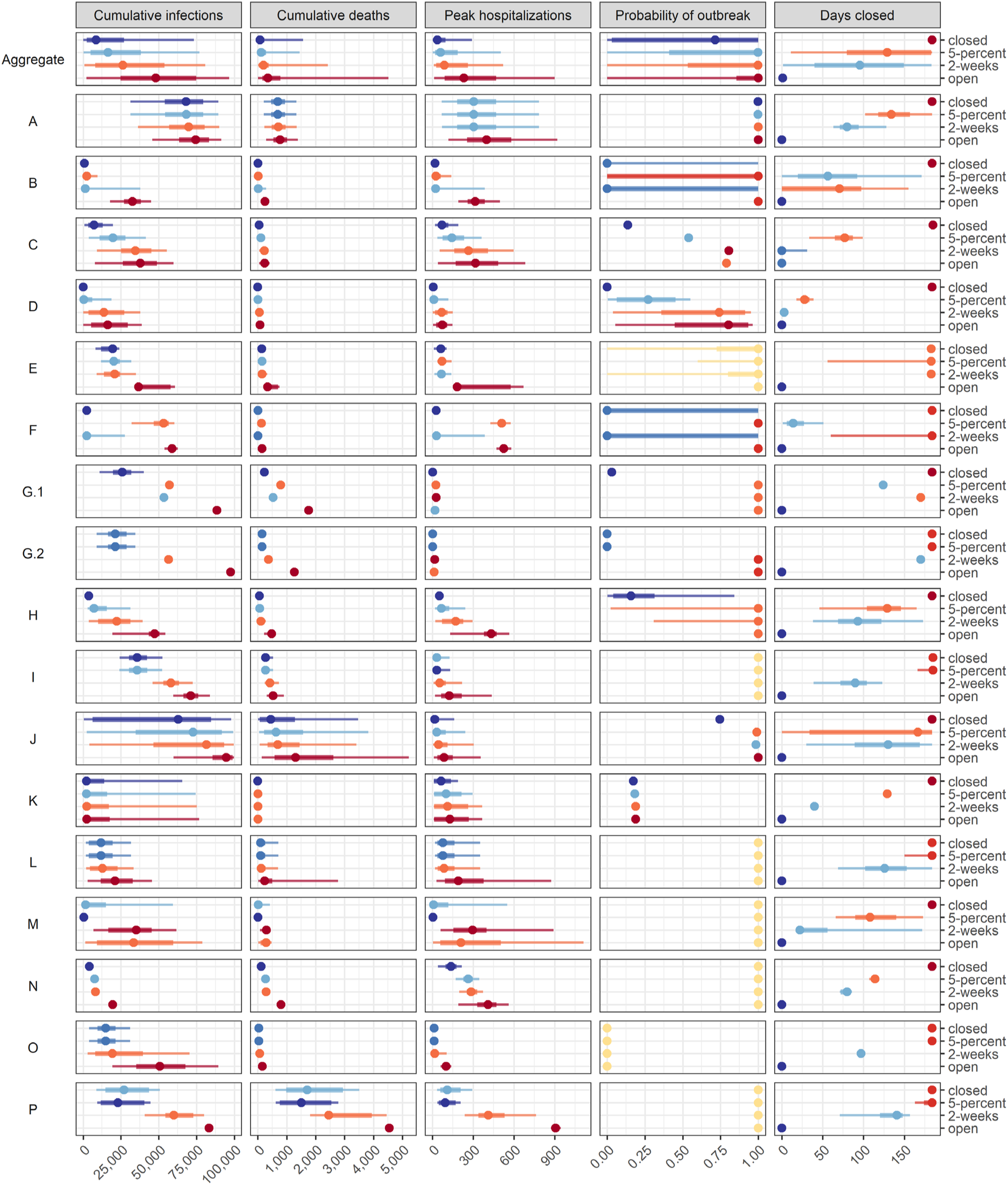
Model results for each target objective and intervention scenario pair organized by model. Median, 50% prediction interval (PI), and 90% PI are indicated as points, thick lines, and thin lines, respectively. Colors denote ranking of each intervention by model for a single objective, where dark blue signifies the lowest value (best performance) and dark red signifies the highest value (worst performance). Ties in ranks are colored as intermediate values. A tie between ranks 1 and 2 and ranks 3 and 4 are shown as an intermediate blue and red, respectively; yellow indicates a tie in ranks across all interventions. Each group is assigned a random, unique identification letter that is specified on the vertical axis.

**SM Fig 2:**
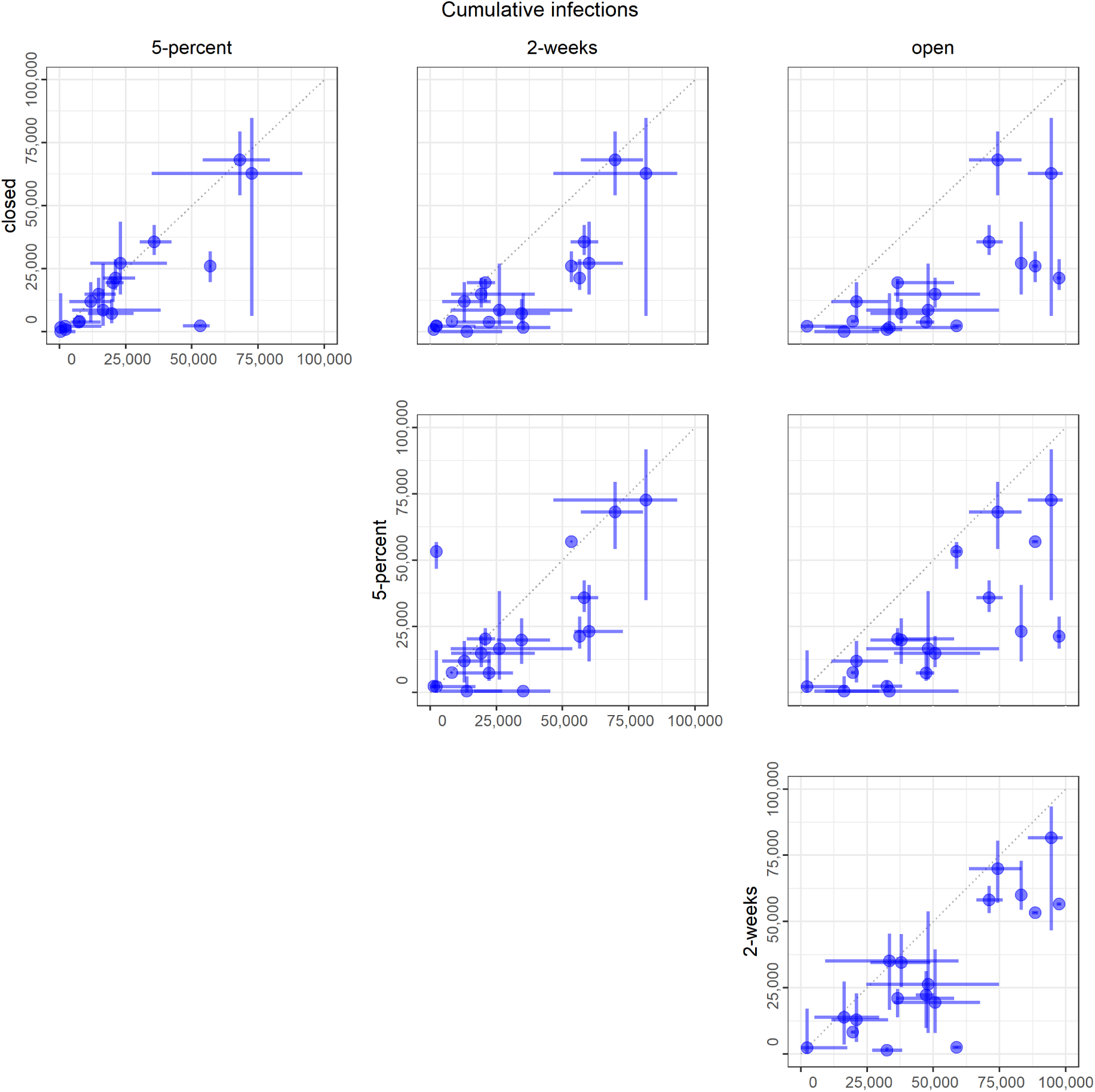
Cumulative infections. Medians (points) and 50% PIs (lines) displayed pairwise by intervention scenario. Each point represents one model.

**SM Fig 3:**
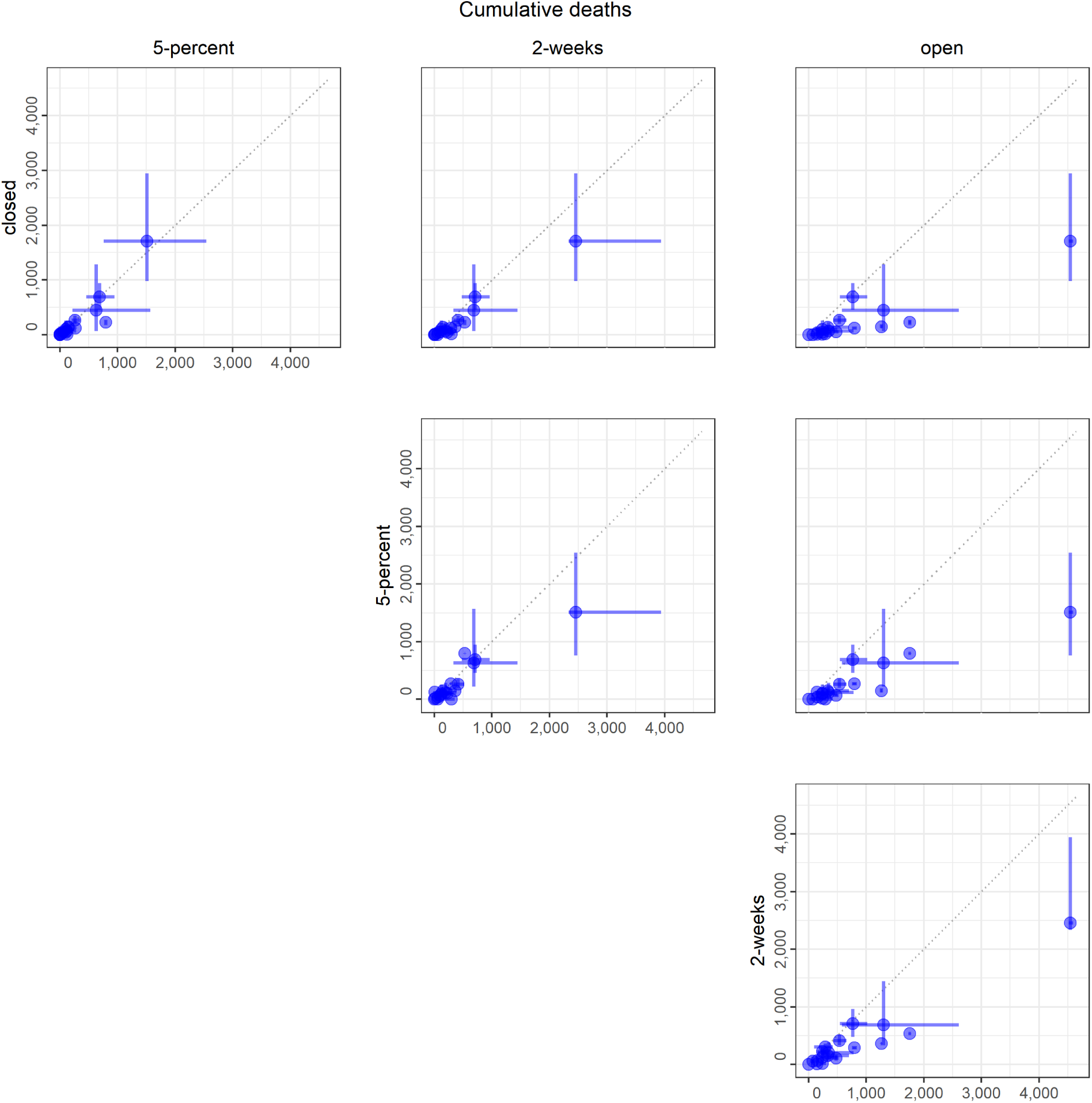
Cumulative deaths. Medians (points) and 50% PIs (lines) displayed pairwise by intervention scenario. Each point represents one model.

**SM Fig 4:**
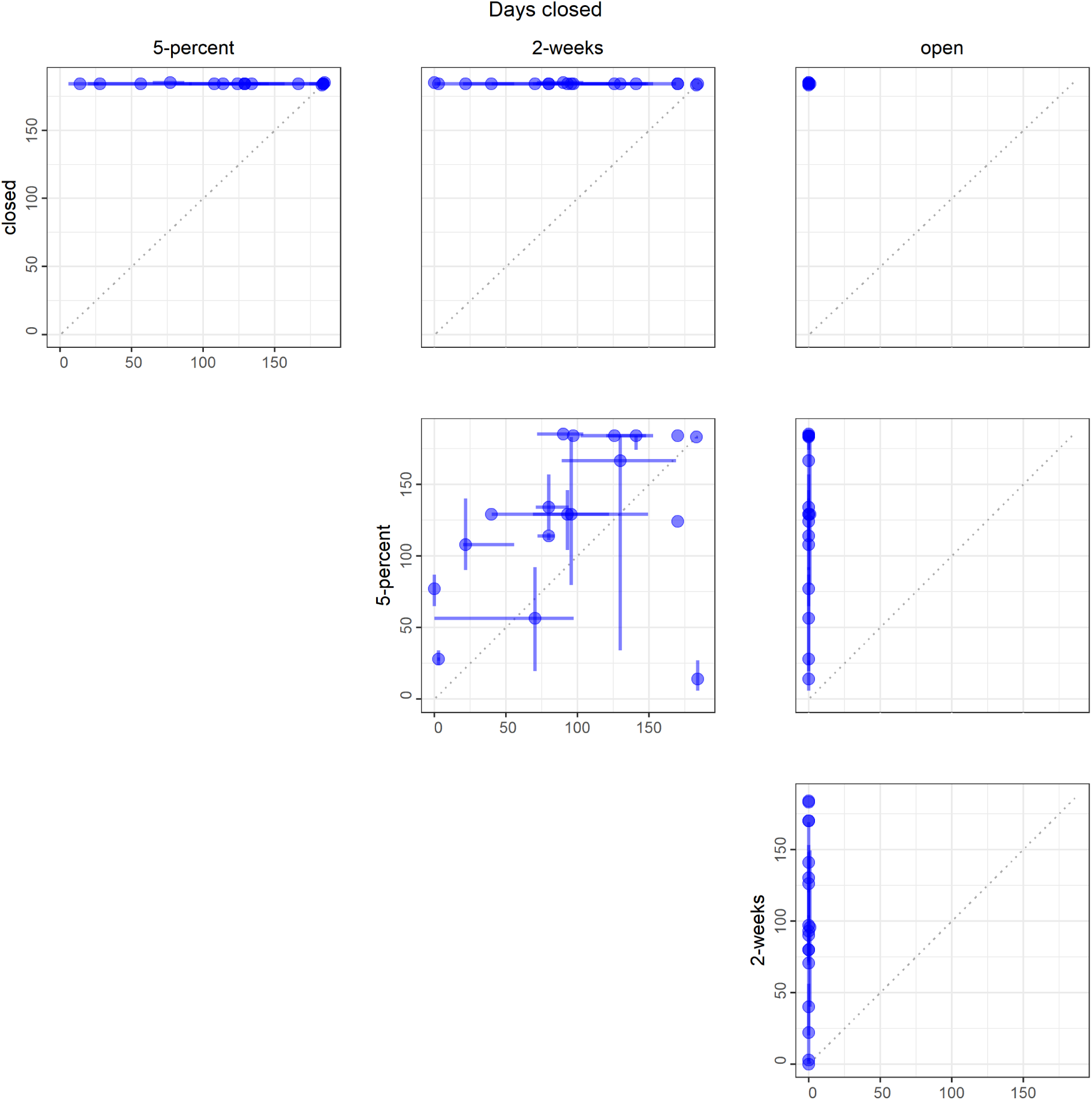
Days closed for non-essential workplaces. Medians (points) and 50% PIs (lines) displayed pairwise by intervention scenario. Each point represents one model.

**SM Fig 5:**
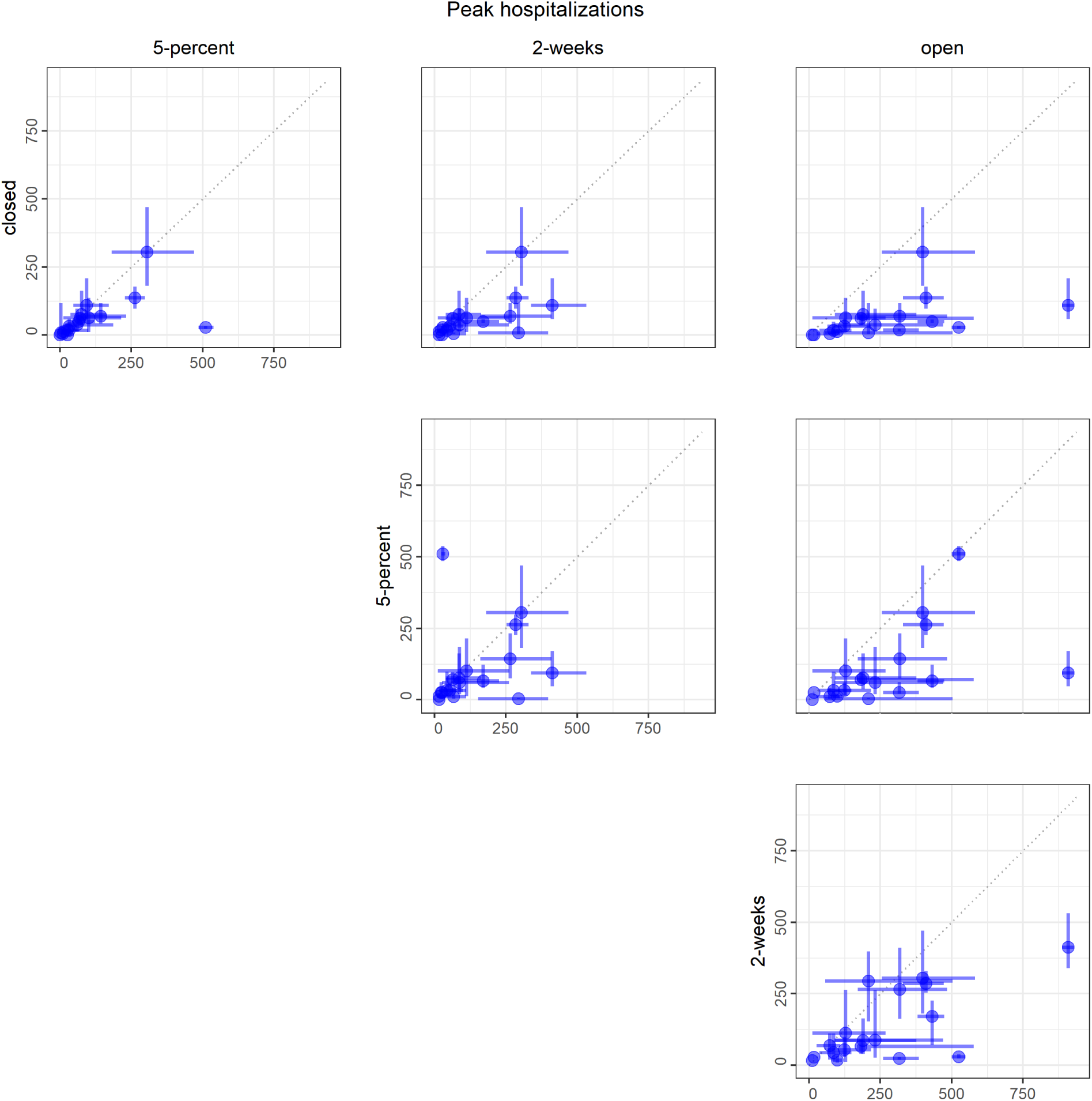
Peak number of hospitalized cases. Medians (points) and 50% PIs (lines) displayed pairwise by intervention scenario. Each point represents one model.

**SM Fig 6:**
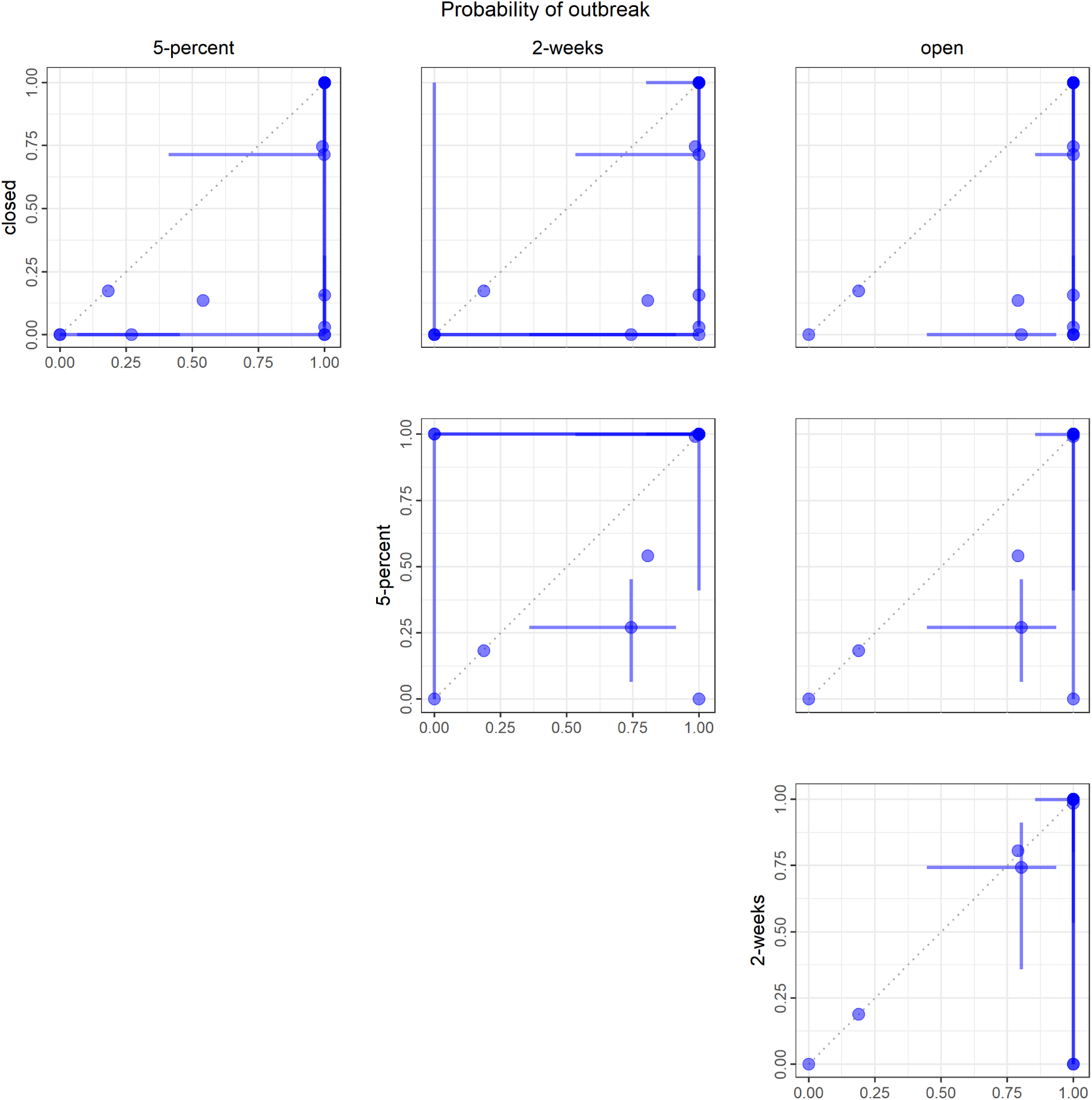
Probability of outbreak. Median (points) and 50% PI (lines) displayed pairwise by intervention scenario. Each point represents one model.

**SM Fig 7:**
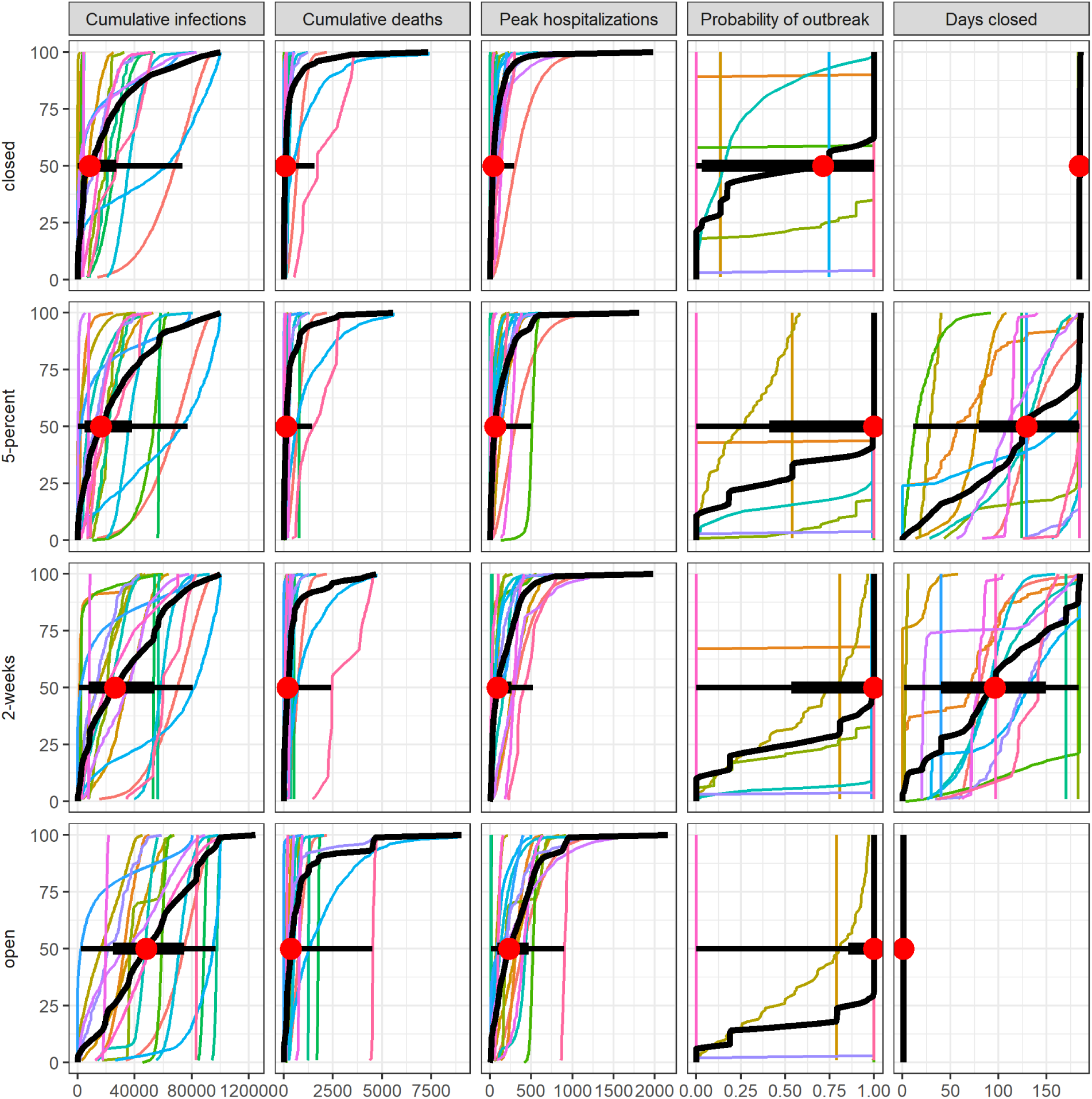
Cumulative distribution functions (CDFs) across models and for the aggregate. Each colored line shows the quantile distribution for a single model. The aggregate CDF is shown in black with median, 50% PI, and 90% PI indicated as red points, thick lines, and thin lines respectively.

**SM Fig 8:**
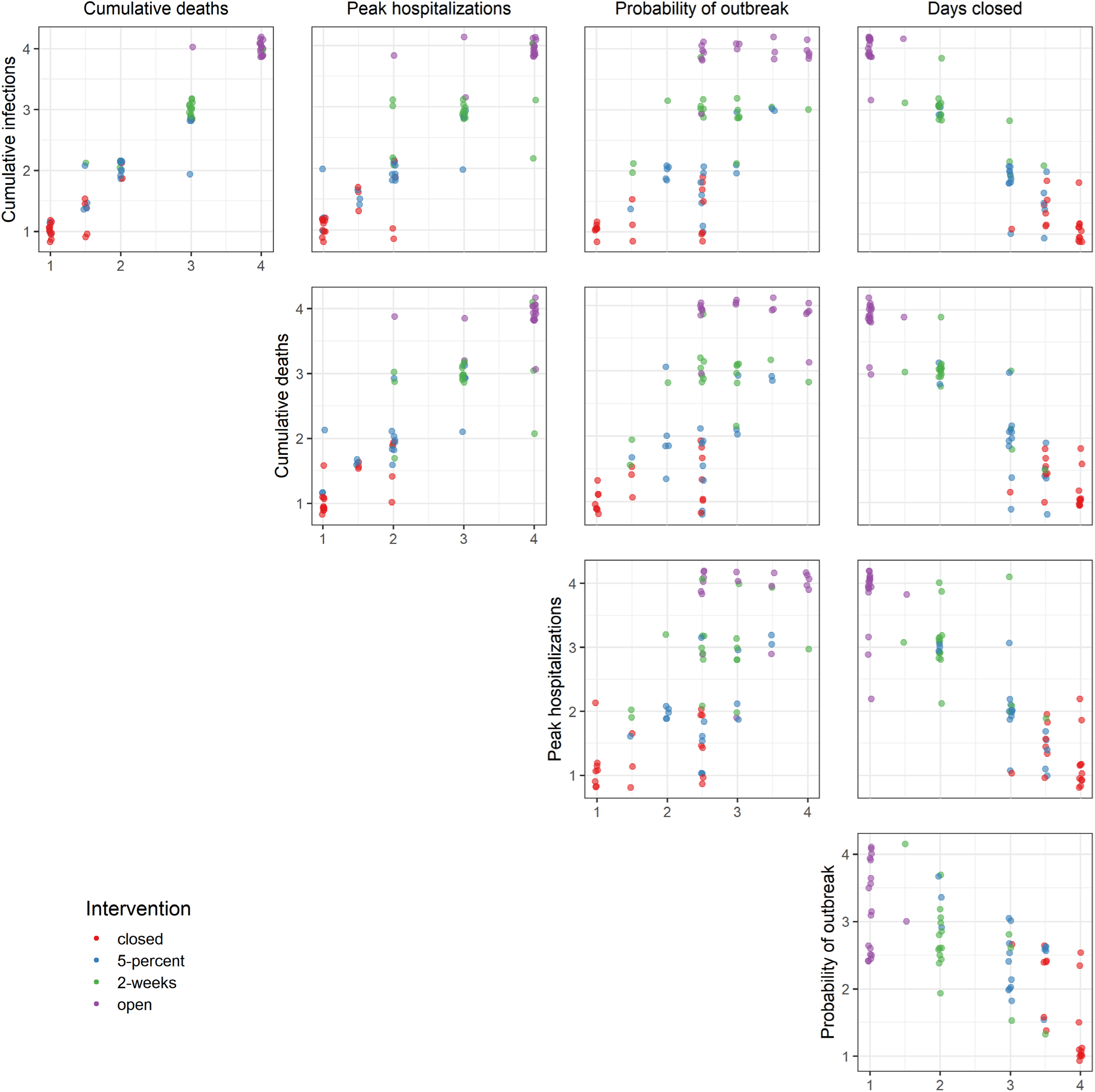
Scatter plots of intervention ranks for a given pair of objectives. Rank ties are shown as intermediate numerical values (e.g. a tie between 1 and 2 is shown as 1.5). For visual clarity, shaded points are jittered around the discrete rank values.

### Supplementary Material: Resolution of linguistic uncertainty in structured discussion between rounds 1 and 2

The group discussion between modeling rounds is an efficient method to reduce linguistic uncertainty resulting from differing interpretations of the problem setting. As well as allowing a common definition of “peak” and other terms, as described in the main text, other sources of unanticipated uncertainty were resolved. For example, one modeling group asked for clarification on the definition of ‘death.’ There was a thorough discussion of the options that different groups had considered or used (reported only; reported plus probable; reported, probable and co-morbidities; or, also indirect deaths, such as those from unrelated causes in patients choosing not to go to the ER during a pandemic). We agreed as a group to use all deaths due to COVID-19 disease-induced mortality, regardless of reporting. This way of counting deaths is based on infection status, not testing status, and can include comorbidities but not indirect deaths, as we are only focusing on people who have been infected with SARS-CoV-2 and died from their infection.

The first round also provided some important checks and balances on the consistency of objective and intervention interpretations across groups, i.e., were the same definitions of workplace closures used? In the first round, some groups used the May 15 to Nov 15 timeframe, others based start dates on declarations of a State of Emergency or stay-at-home orders, and one group implemented a weighting for essential and non-essential business closures and associated compliance issues explicitly (SM Fig. 9). Including a metric that should be consistent across models allowed us to check for and remove linguistic uncertainty in round 2 submissions that would have limited our ability to compare the rankings of interventions between models and objectives.

In addition to resolving linguistic uncertainty, the first round provided information on the utility of the interventions themselves. We initially requested results for reopening after declining to 1% of peak – round 1 results suggested this condition would rarely, if ever, be met, and thus we altered the intervention to trigger at 5% of peak, instead. Typically, such changes in interventions would be made in consultation with decision makers (as part of Fig. 1, loop A).

Deliberately, consensus on scientific uncertainty was not required. In fact, model results were presented anonymously to reduce the pressure to conform to other groups’ expectations and hence to avoid ‘groupthink,’ and other cognitive biases, engendering a more comprehensive expression of legitimate scientific uncertainty. We thus encouraged modeling groups to adjust their models to reflect unknown aspects of the transmission and intervention implementation process to more fully express genuine scientific and logistical uncertainty.

Due to the opposing effects of decreasing linguistic uncertainty and maintaining or increasing expression of scientific uncertainty, it was difficult to draw conclusions about the source of model-level changes in expressions of uncertainty between rounds 1 and 2. To begin to assess model-level changes, we compared the lengths of inter-quartile ranges (IQRs) (SM Figs 10-12) within groups by round as well as the ratio of IQR length between each model and the corresponding aggregate distribution. The clearest examples of model incorporation of additional scientific uncertainty in round 2 were the models that provided point estimates in round 1 (length of IQR = 0) that subsequently expanded these estimates to distributions in round 2. Requiring distributions rather than point estimates necessarily increased the degree of expressed uncertainty. However, even in these models, we observed decreases in uncertainty (presumably in linguistic uncertainty) as the point estimates account for the majority of outliers in round 1 (SM Fig. 10).

For each objective-intervention pair considered in both rounds, the length of the aggregate IQR was greater than the median length of the corresponding model IQRs (SM Fig. 11). The degree of uncertainty (as measured by IQR lengths) for the majority of models increased towards that of the corresponding objective-intervention aggregate distribution from round 1 to round 2 (see the clustering of points near the orange dashed line in SM Fig. 11 round 2).

Implementation of the open and closed interventions did not rely on a definition of “peak”. In SM Fig. 12, we observed that the ratio of IQRs (IQR(model)/IQR(aggregate)) between rounds tended to be closer to one than the 2-week intervention, which required a definition of peak (SM Fig. 12). We also note that decreases in the IQR length for the aggregate distribution were observed for all objectives in the 2-week scenario (i.e. aggregate ratio of IQR _R2_/IQR _R1_ <1). Changes observed in the open scenario (cumulative infections, cumulative deaths, and peak hospitalization ratios observed are 1.20, 1.02, and 0.949 respectively) were moderate compared to those in the closed scenario (cumulative infections, cumulative deaths, and peak hospitalization ratios observed are 1.93, 1.92, and 1.54 respectively). Note that an analogous comparison for the alternative peak intervention was not possible, given the change from 1-percent of the peak to 5-percent of the peak between rounds.

**SM Fig 9:**
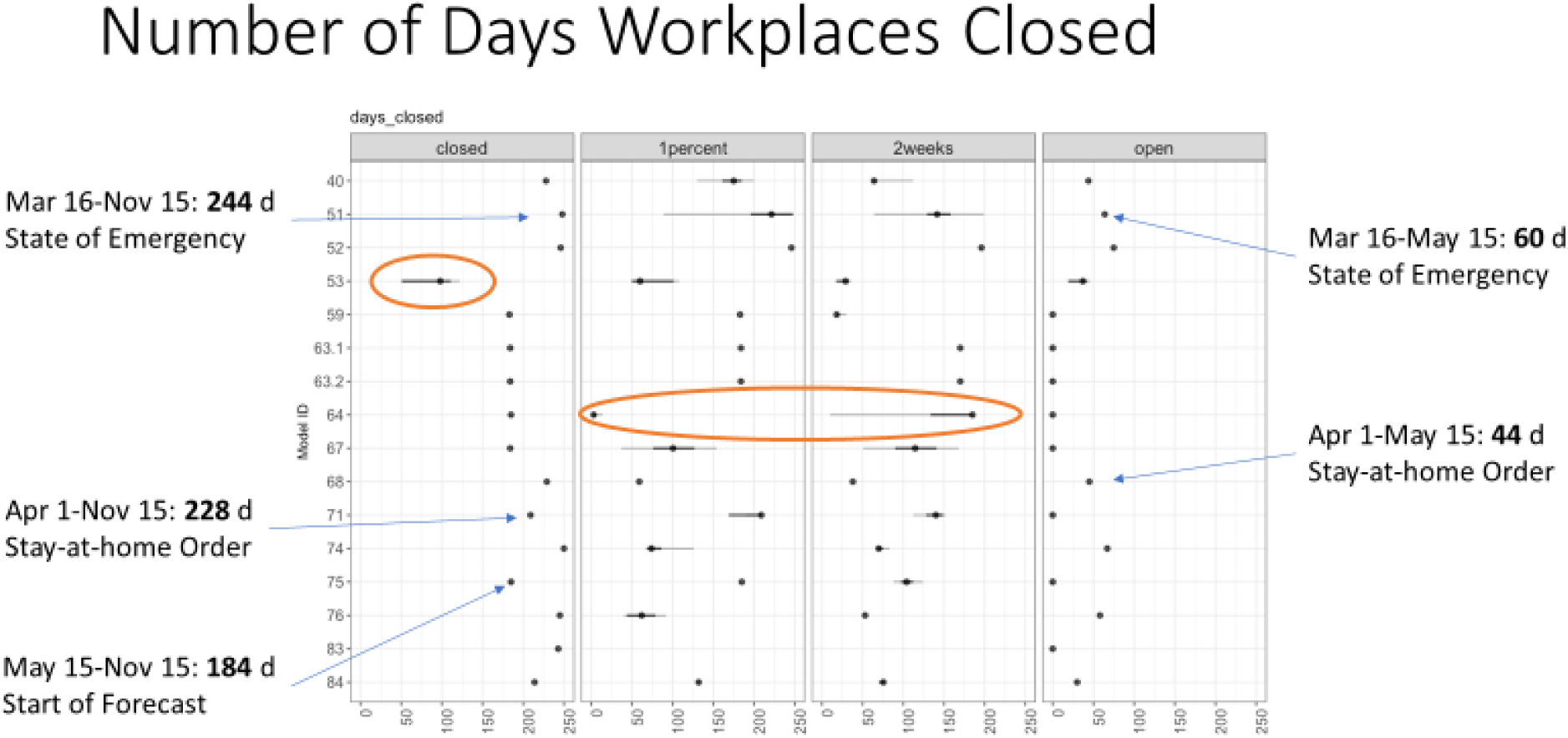
Resolution of linguistic uncertainty about the number of days non-essential workplaces are closed in the discussion following round 1 of modeling (note that model IDs changed between rounds); figure is of a slide from the group discussion after round 1. See main text Fig. 3 column 5 for days of non-essential workplace closure results from round 2. Ovals highlight points of discussion about different ways of capturing uncertainty for days workplaces are closed and unusual results about intermediate interventions.

**SM Fig 10:**
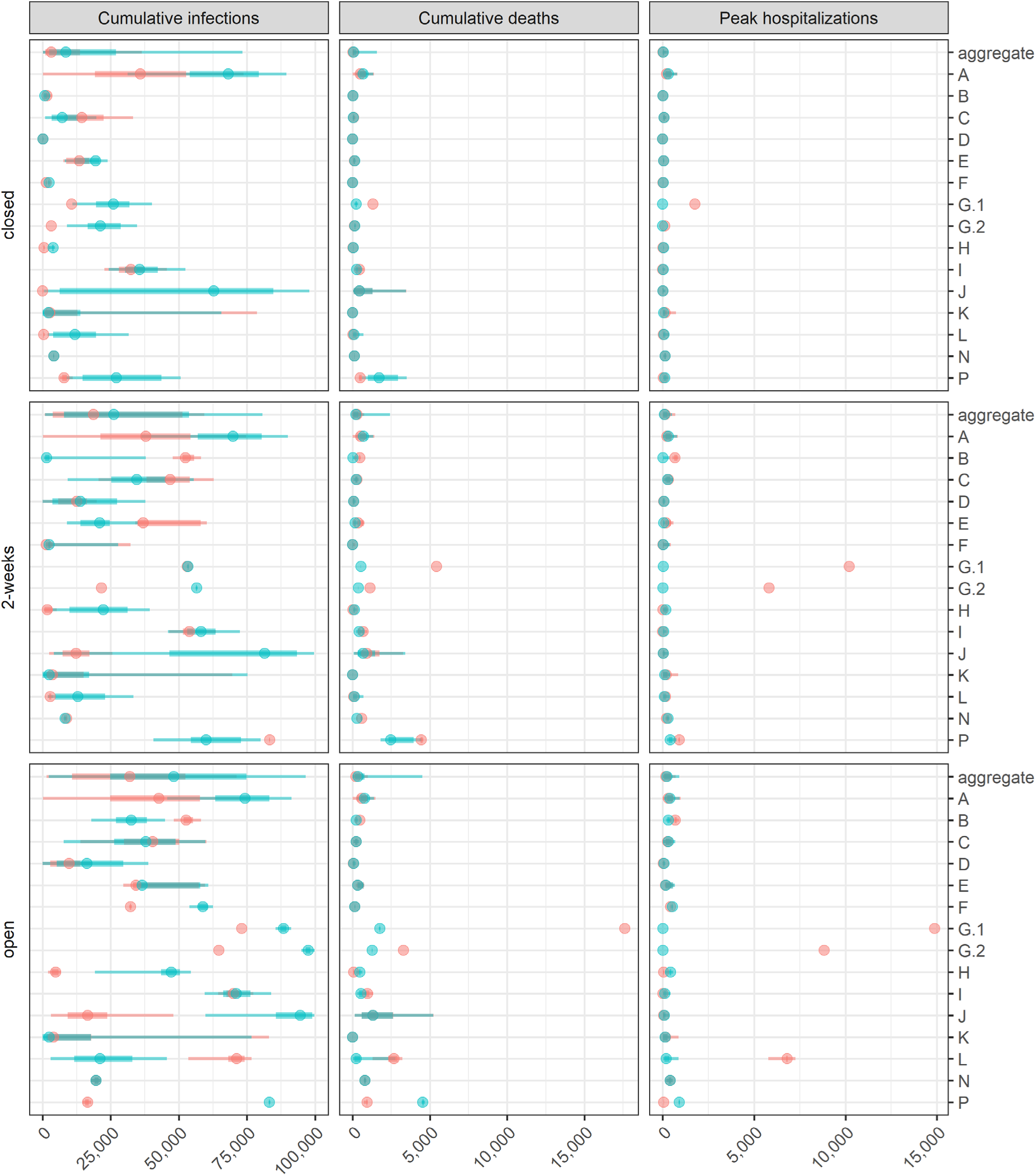
Team and aggregate values for each intervention and objective pair. Round 1 and round 2 results displayed in red and blue respectively. Since the 1-percent intervention from round 1 was updated to a 5-percent intervention in round 2, results for these interventions have been omitted from this comparison. Also note that two models were excluded from this analysis, as they submitted incomplete results in round 1. After the discussion between rounds 1 and 2, these groups were able to provide complete and comparable results. Additionally, in at least one case, some of the differences can be attributed to model error fixes between rounds.

**SM Fig 11:**
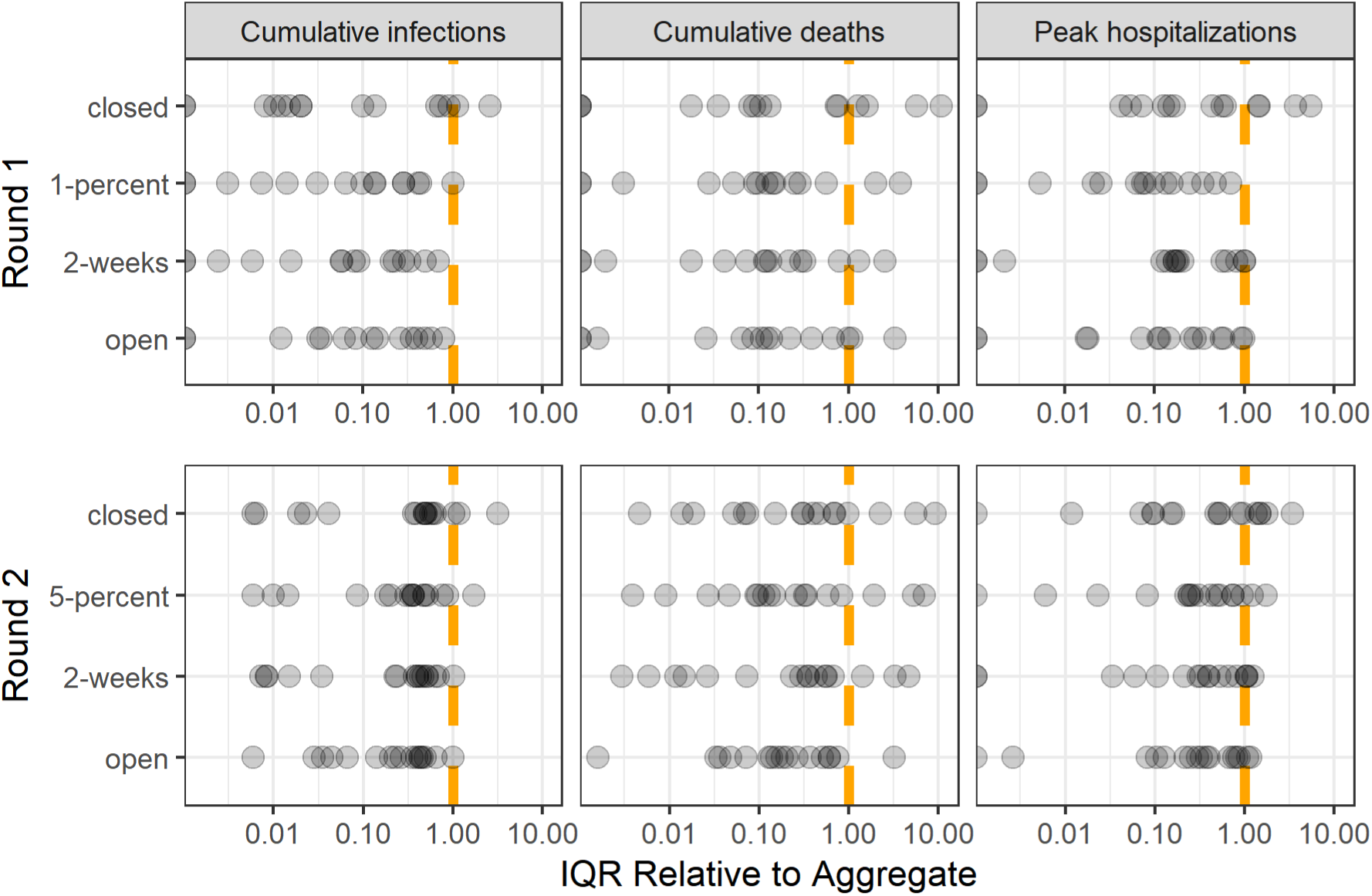
Comparison between model-specific IQR lengths and the length of the IQR for the aggregate distribution (i.e. length(IQR_team)/length(IQR_aggregate)) shown on a logarithmic scale. Results are grouped by round, intervention, and objective. Round is indicated on the left axis. Columns indicate the objective and rows indicate the intervention. The dashed orange line highlights the point at which there is no difference between the model-specific IQR lengths between rounds 1 and 2 (points to the left indicate a model IQR less than that of the corresponding aggregate and *vice versa* for points to the right).

**SM Fig 12:**
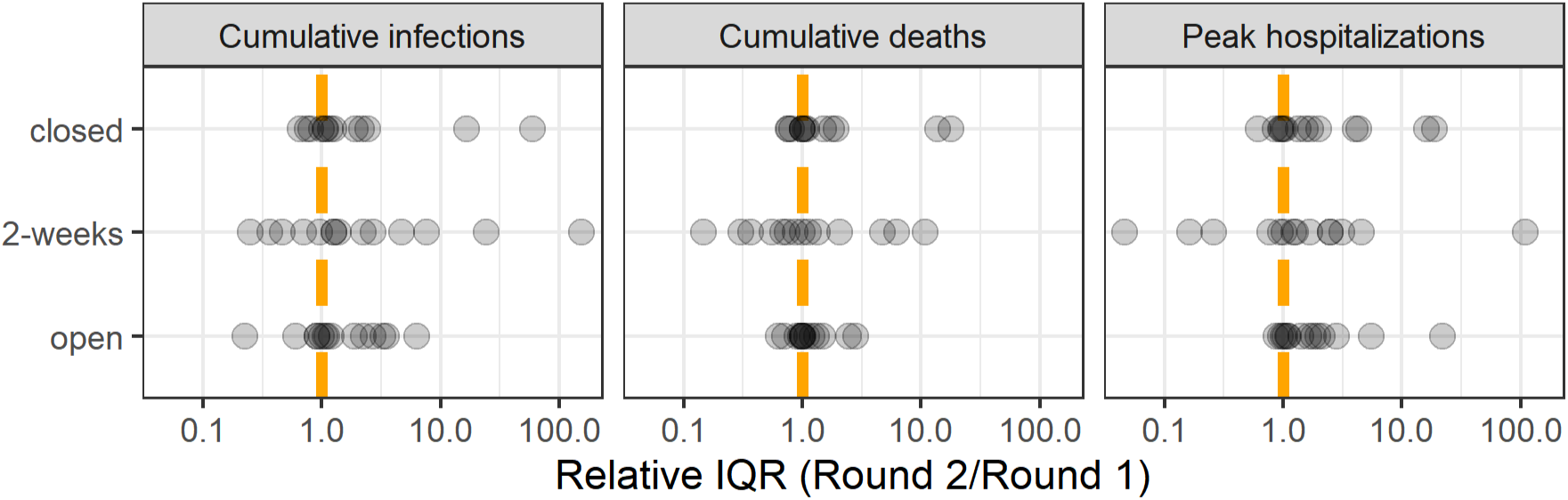
Round comparison of IQR length by team, calculated as the ratio of the length of IQRs between rounds 1 and 2 (i.e. length(IQR _R2_) / length(IQR _R1_)) shown on a logarithmic scale. Note that in the first round, two models (G.1 and G.2) submitted point estimates for each intervention and metric. Since point estimates are such that length(IQR) = 0, the relative IQR (compared to round 1) is infinity and thus not shown here. Similarly, there is not a point representing cumulative deaths in the closed scenario for group K since the corresponding length(IQR) = 0. Because the 1-percent intervention from round 1 was changed to a 5-percent intervention in round 2, the corresponding results have been omitted from this comparison. The dashed orange line highlights the point at which there is no difference between the model-specific IQR lengths between rounds 1 and 2 (points to the left indicate a lower R2 IQR than that of the corresponding group’s R1 submission, and vice versa for points to the right).

### Supplementary Material: Comparison of county data with aggregate model results

The modeling exercise was motivated by a U.S. county representative of mid-sized counties with populations of approximately 100,000 people, with limited mobility and stay at home orders in place until at least May 15, 2020. Here, we compare aggregate model results with reported data from counties meeting the target county description.

We first selected the 99 U.S. counties with population sizes between 90,000 and 110,000 using data from the Johns Hopkins University COVID-19 dashboard (Dong, Du, and Gardner 2020). From this subset, we then selected counties with stay at home orders in place until at least May 15, 2020 (data from Killeen et al. (2020), Keystone Strategy (2020), and NACO county emergency declarations (2020)), and changes in mobility in line with stay at home orders, i.e., less than 50% increase from baseline retail mobility, less than 25% increase in baseline work mobility, and less than 5% decline from baseline residential mobility (data from Google COVID-19 Community Mobility Reports). This resulted in a subset of 84 counties. Finally, from this subset, we determined the set of counties implementing a fully ‘closed’ intervention (with stay at home orders in place from May 15, 2020 to present day (October 15, 2020 as of submission) <u>and</u> mobility patterns suggesting those orders were followed). We found 66 counties that met these criteria. No counties were found to be fully open during this period, and it was not possible to determine if any counties implemented the ‘2-week’ or ‘5-percent’ interventions. We compared aggregate cumulative deaths (reported deaths only) with modeled cumulative deaths (all COVID-19 deaths) under the closed intervention for the 66 counties following the ‘closed’ intervention. Cumulative reported deaths for the 66 counties under the closed intervention were summarized in 100 quantiles, the same format requested from model groups (See SM Figs 13 - 14, below).

Note that model results were for the entire period May 15, 2020 to November 15, 2020 and data were only available through the present day (October 15, 2020 as of submission). Further, we are comparing *reported* deaths (from data) with *all* COVID-19 deaths, not just reported deaths (from model results). We did not assume a reporting rate, but expect a higher number of model predicted cumulative deaths. Crucially, our results represent the realization of one pandemic across 66 counties in comparison to multiple model realizations across a wide range of uncertainty. Thus, the model uncertainty will necessarily be higher than the observed uncertainty. The model mean will likely also be higher, as the right-skewed uncertainty will increase the mean.

County comparison results provided in the SM will be updated once data are available for the entire period May 15, 2020 to November 15, 2020, for counties that remain closed.

**SM Fig 13:**
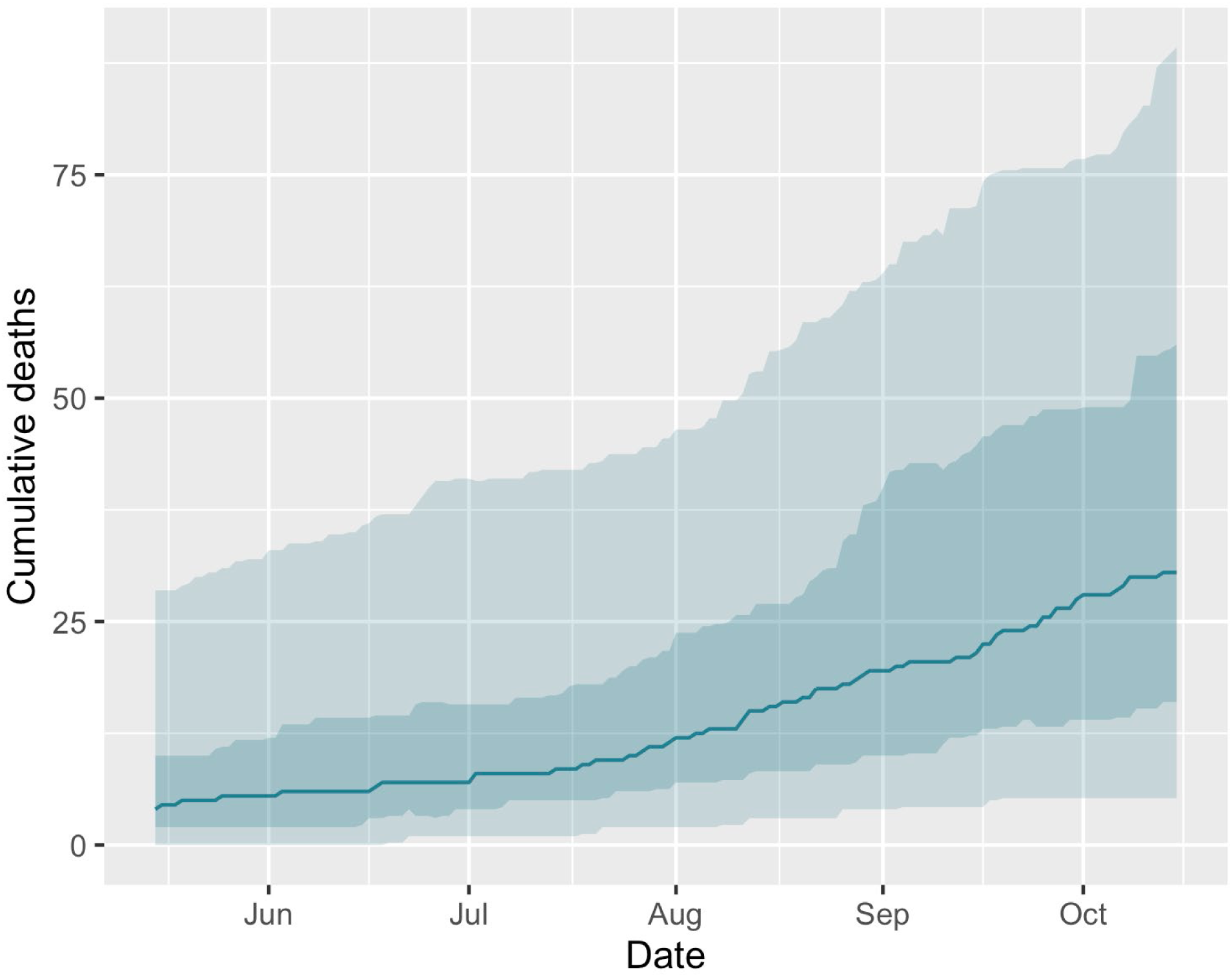
Summary of cumulative reported deaths for counties similar to the model context and following the closed intervention. Median reported cumulative deaths (solid line), 50% IQR (darker shaded area), and 90% IQR (lighter shaded area) for the subset of 66 counties following the closed intervention from May 15 to October 15.

**SM Fig 14:**
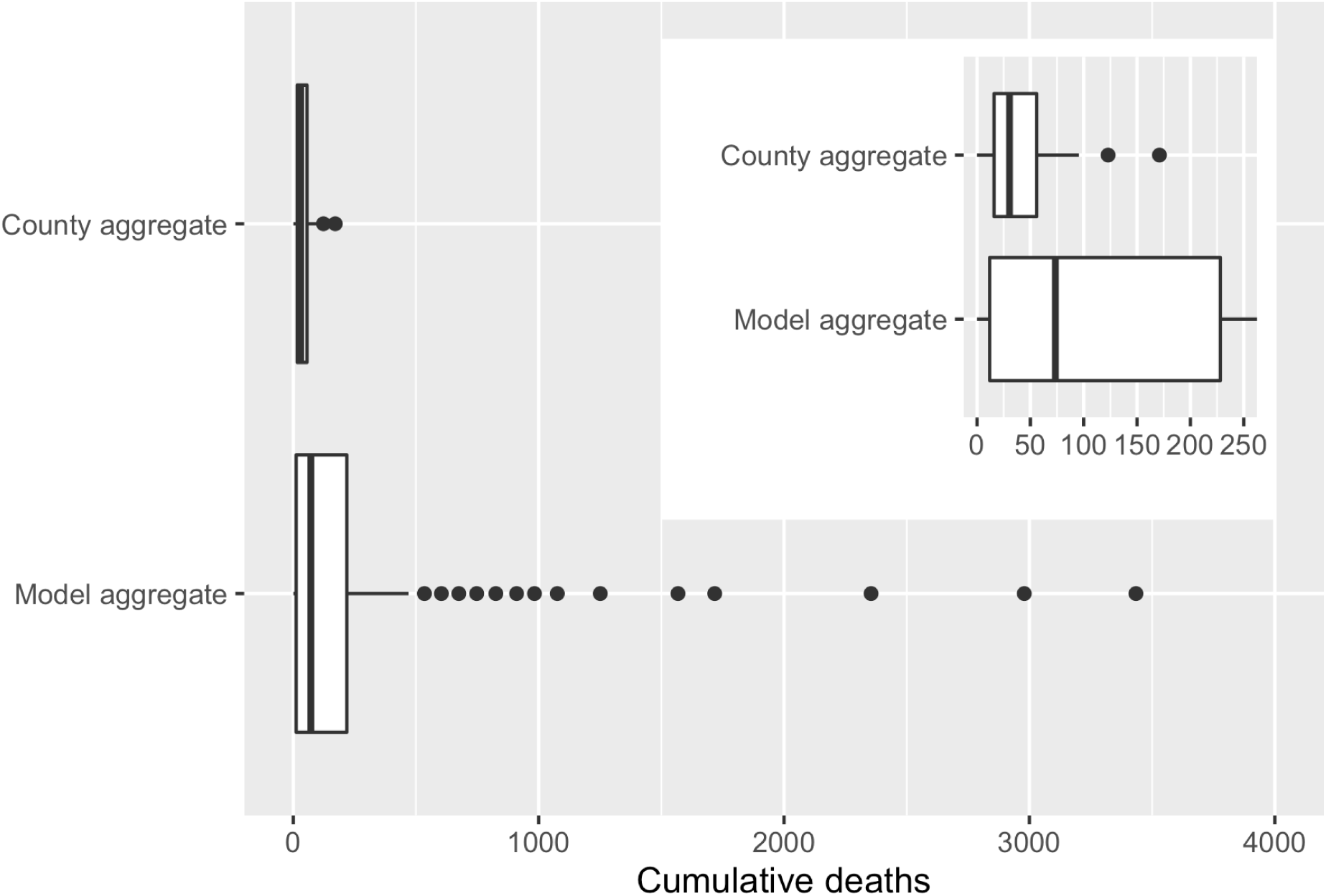
Comparison of aggregate reported county data to model results for the closed intervention. Boxplot of cumulative reported deaths from 66 U.S. counties from May 15 to October 15 (median: 31; 50% IQR: 16, 56; 90% IQR: 3, 59) and model results for cumulative deaths from May 15 to November 15 (median: 73; 50% IQR: 12, 228; 90% IQR: 2, 1568). Inset shows overlap of box area for the two plots.

## Supplementary Material: Checklist Data

**SM Table 1:**
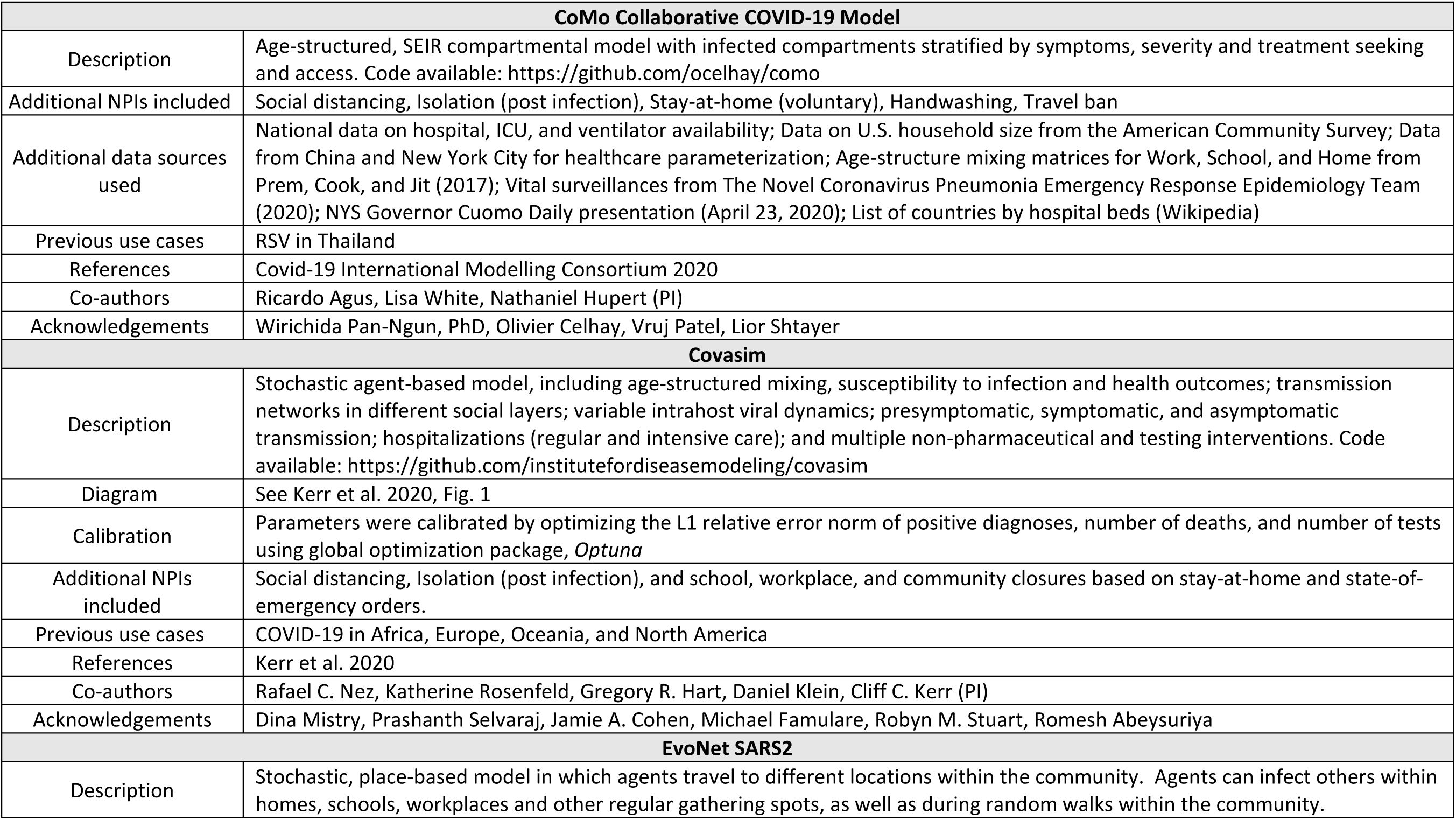
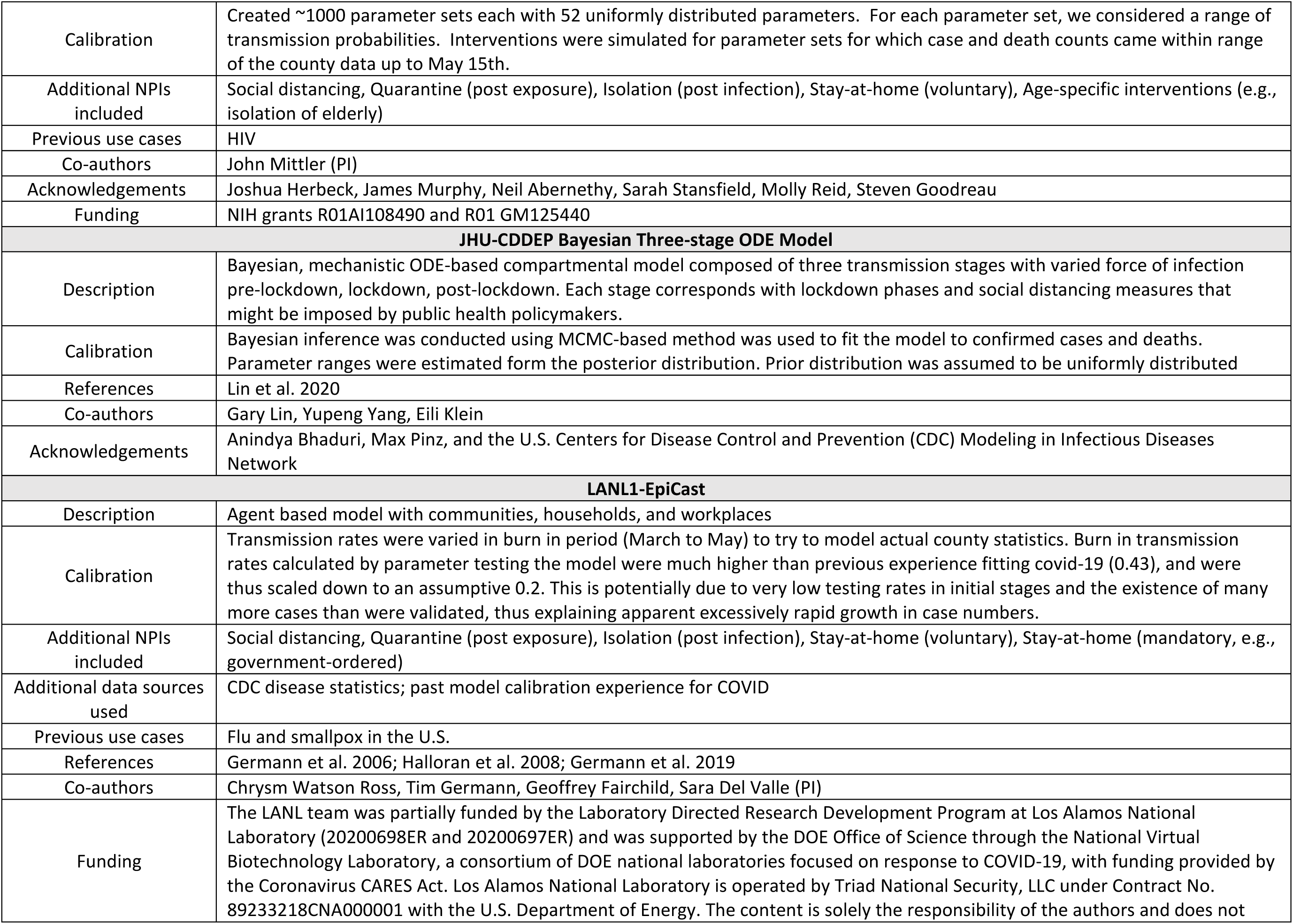
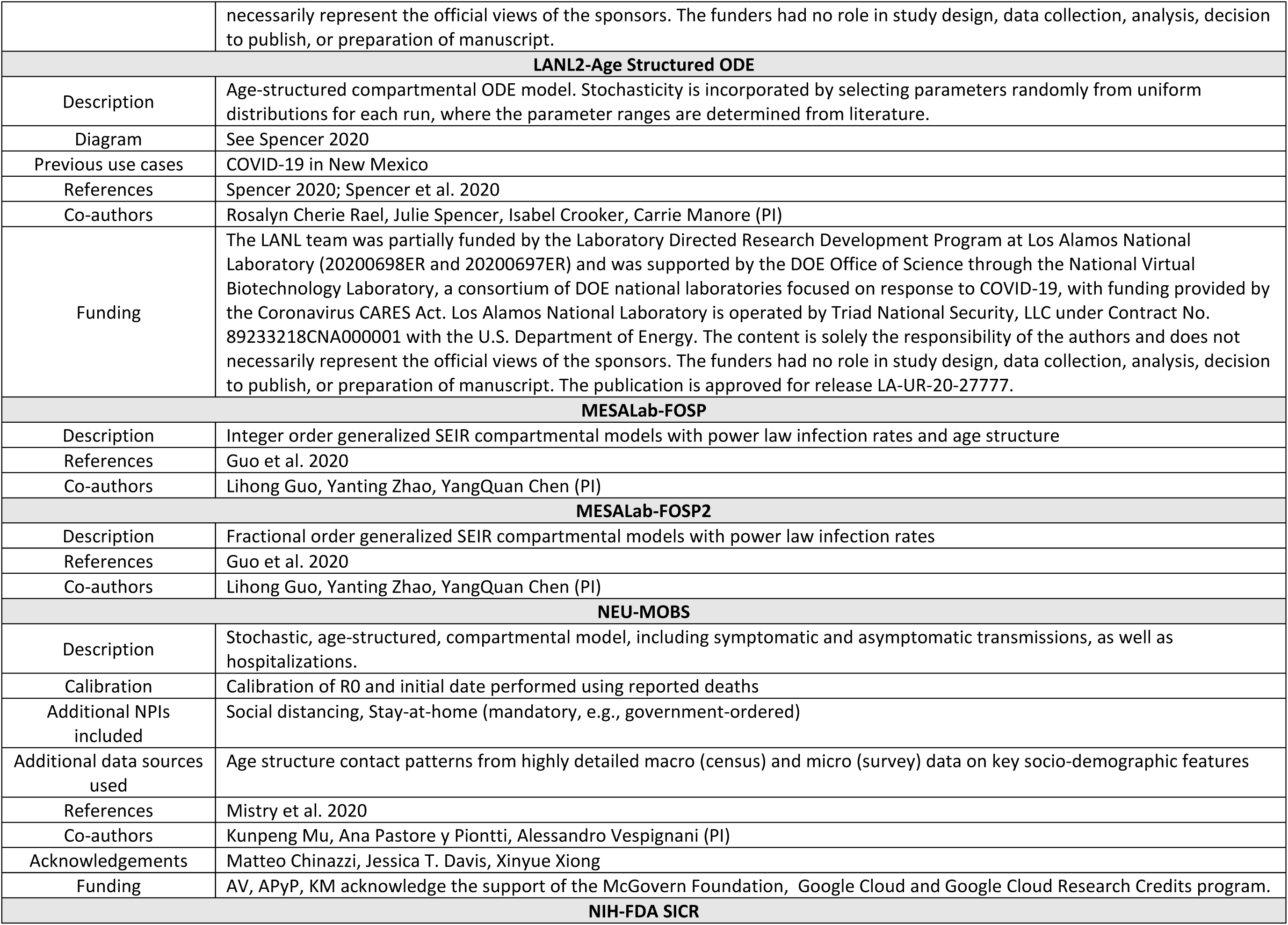
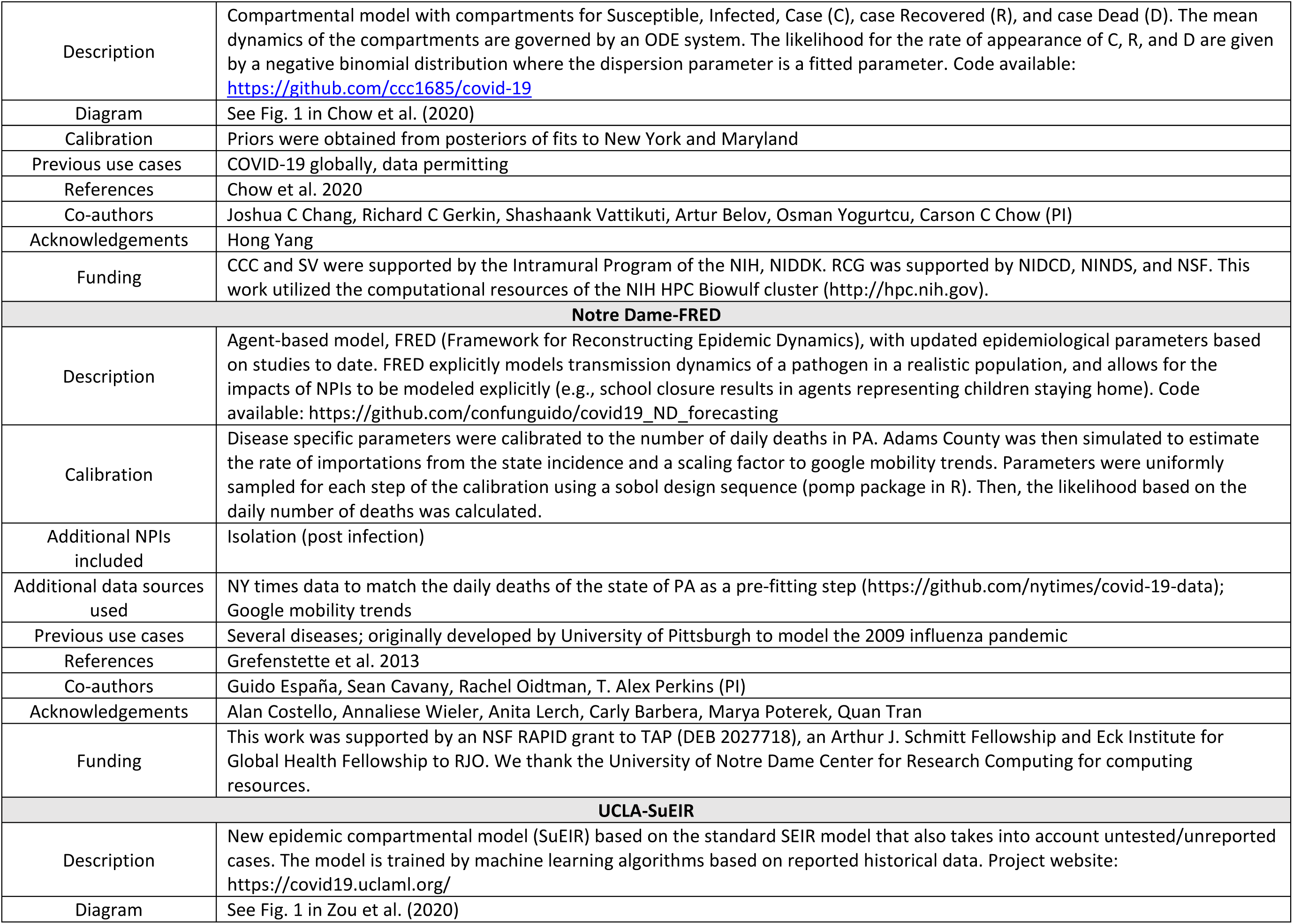
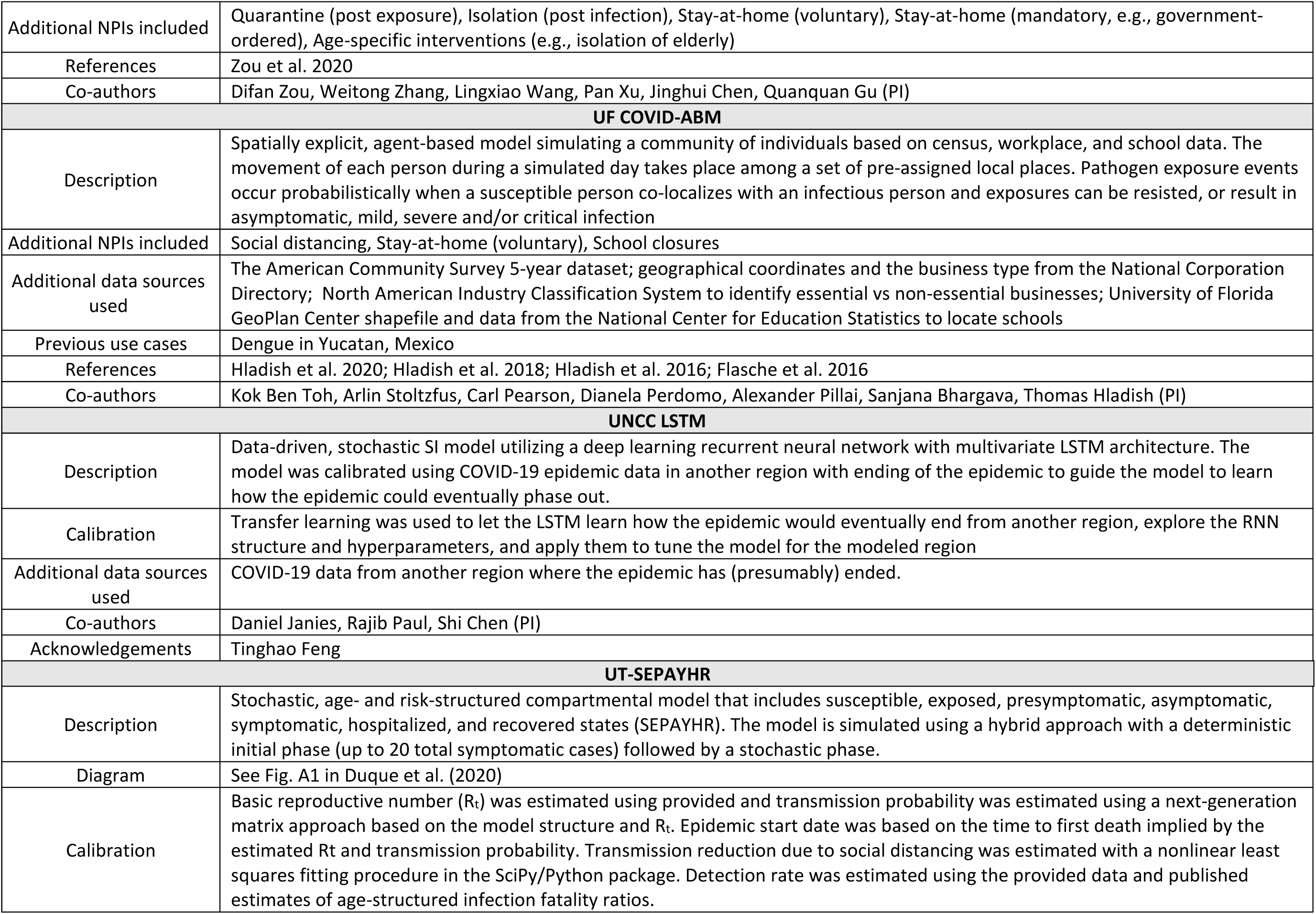
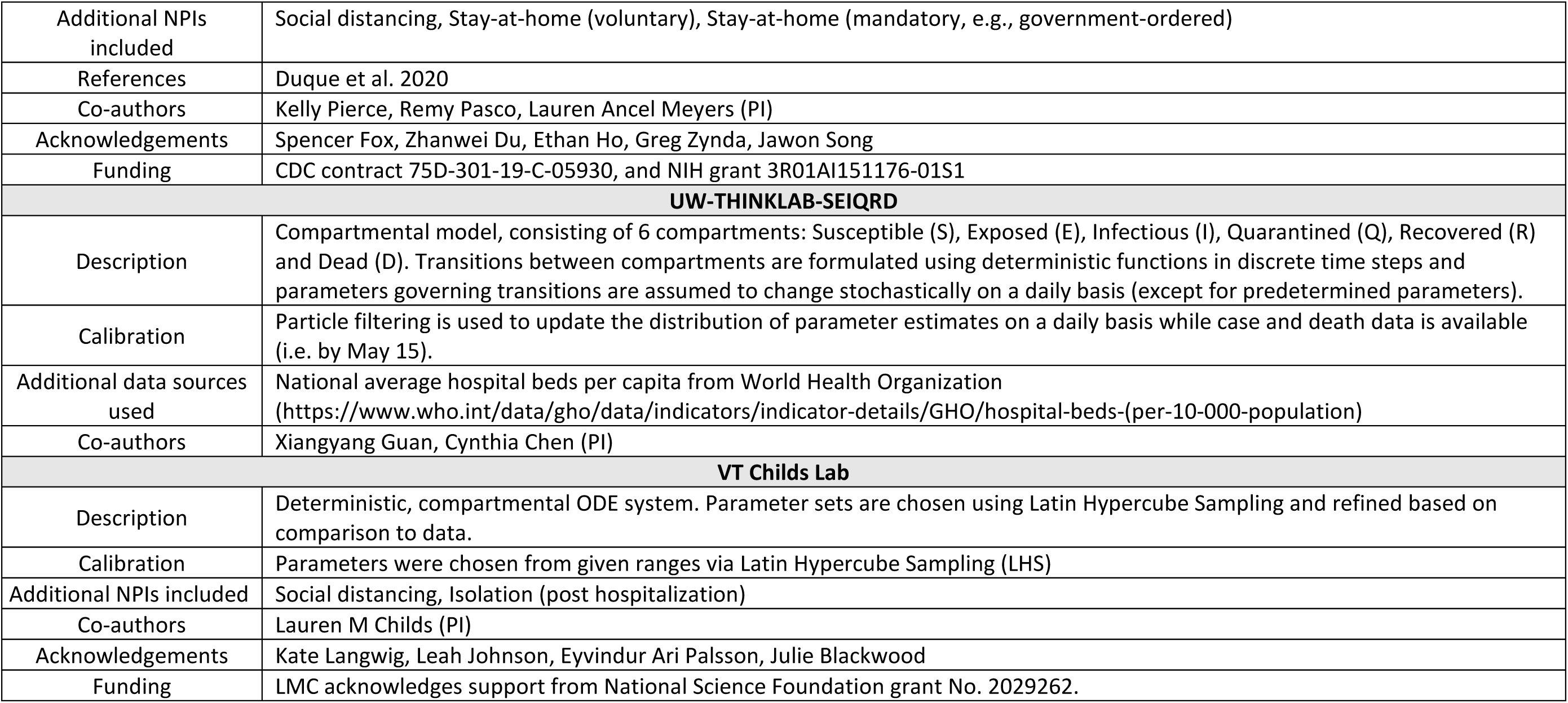
Contributed model descriptions. Name, description (including links to model code where available), diagram, calibration method, other non-pharmaceutical interventions (NPIs) included in the model, additional data sources used, previous use cases for the model (both for COVID-19 in other settings and other disease systems) and references for each of the 17 models. Categories which were not relevant were excluded.

**SM Table 2:** Importation rate. Most models did not include an importation rate after any initial seeding. Models that did maintained a relatively small importation rate, per the elicitation setting.

| Model ID | Importation rate (or None) |
| --- | --- |
| A | None |
| B | None |
| C | None |
| D | None |
| E | 0.14 cases / day |
| F | 1 exposure / day (with a probability of resistance) |
| G.1 | None |
| G.2 | None |
| H | None |
| I | None (after initial seeding) |
| J | None (after initial seeding) |
| K | None |
| L | 0 – 0.5 cases / day |
| M | None |
| N | None |
| O | Varied |
| P | None |

**SM Fig 15:**
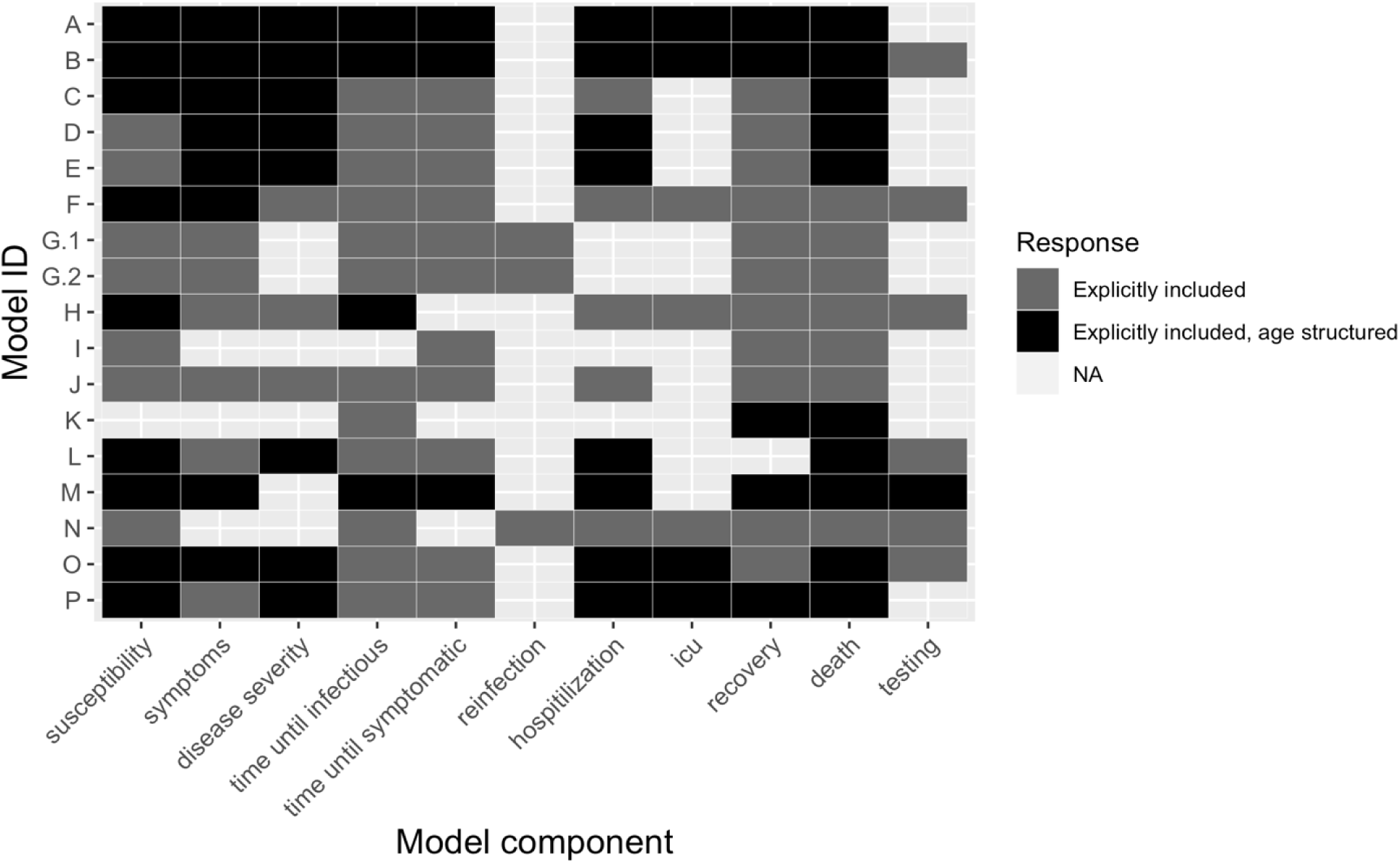
Description of model components and structure by model. Participants were asked to indicate which model components were included in their model (from a given set) and whether any component was structured by age and/or gender and/or sex as part of the submission checklist. No model included any components structure by gender and/or sex. Twelve of the 17 included at least one component structured by age.

**SM Fig 16:**
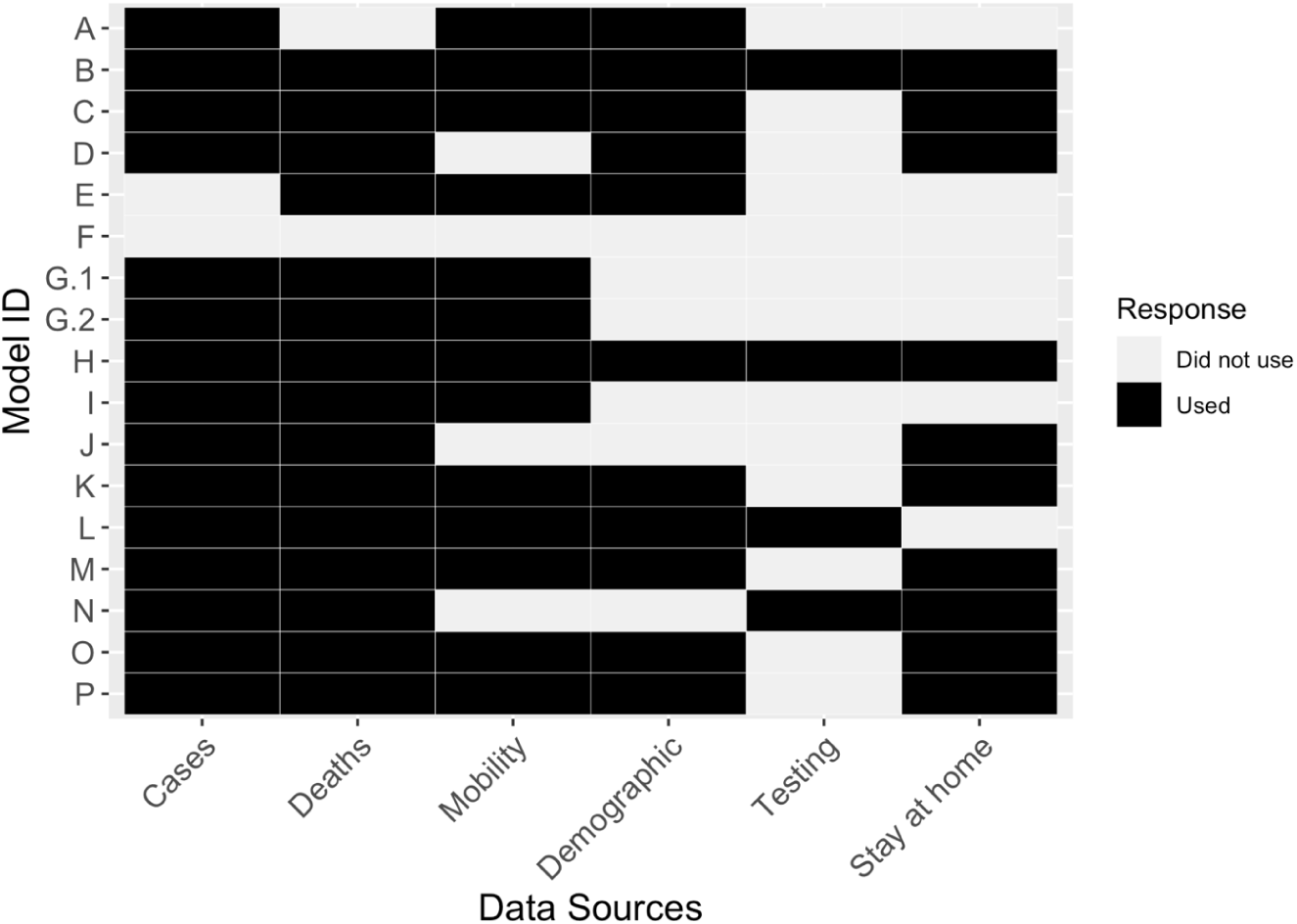
Data sources used for each model. Participants were asked to indicate which of the provided datasets were used for any part of the model (e.g., for calibration, training, fitting etc.) as part of the submission checklist. All but one model used at least two of the provided data. Model F used only external data sources (provided data was used solely to better understand the intent of the exercise).

**SM Fig 17:**
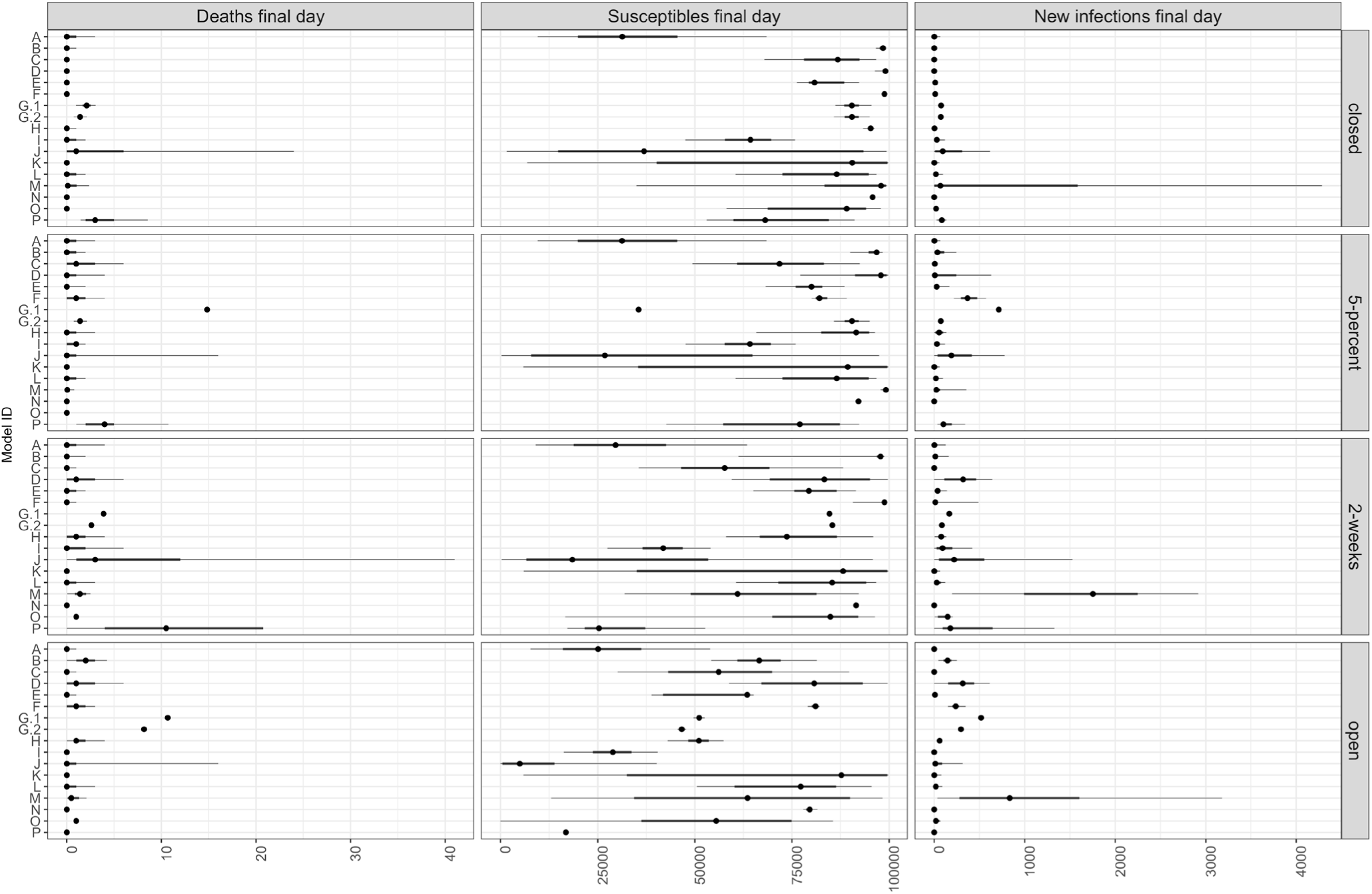
Projected number of deaths, people who are susceptible, and new infections under each scenario for the final day of the forecast. Participants reported the 5 ^th^, 25 ^th^, 50 ^th^, 75 ^th^, and 95 ^th^ quantiles for the number of deaths, susceptibles, and new infections on the final day (November 15, 2020) under each scenario. All models started with similar initial susceptibles.

